# Disruption of long-term psychological distress trajectories during the COVID-19 pandemic: evidence from three British birth cohorts

**DOI:** 10.1101/2022.04.22.22274164

**Authors:** Darío Moreno-Agostino, Helen L. Fisher, Alissa Goodman, Stephani L Hatch, Craig Morgan, Marcus Richards, Jayati Das-Munshi, George B. Ploubidis

## Abstract

**Importance:** Mental health disorders were among the leading global contributors to years lived with disability prior to the COVID-19 pandemic onset, and growing evidence suggests that population mental health outcomes have worsened since the pandemic started. The extent that these changes have altered common age-related trends in psychological distress, where distress typically rises until mid-life and then falls in both sexes, is unknown.

**Objective:** To analyse whether long-term pre-pandemic psychological distress trajectories have altered during the pandemic, and whether these changes have been different across generations and by sex.

**Design:** Cross-cohort study with prospective data collection over a 40-year period (earliest time point: 1981; latest time point: February/March 2021).

**Setting:** Population-based (adult general population), Great Britain.

**Participants:** Members of three nationally representative birth cohorts which comprised all people born in Great Britain in a single week of 1946, 1958, or 1970, and who participated in at least one of the data collection waves conducted after the start of the pandemic (40.6%, 42.8%, 39.4%, respectively).

**Exposure(s):** Time, COVID-19 pandemic.

**Main Outcome(s) and Measure(s):** Psychological distress factor scores, as measured by validated self-reported questionnaires.

**Results:** 16,389 participants (2,175 from the 1946 birth cohort, 52.8% women; 7,446 from the 1958 birth cohort, 52.4% women; and 6,768 from the 1970 birth cohort, 56.2% women) participated in the study. By September/October 2020, psychological distress levels had reached or exceeded the levels of the peak in the pre-pandemic life-course trajectories, with larger increases in younger cohorts: Standardised Mean Differences (SMD) and 95% confidence intervals (CIs) of -0.02 [-0.07, 0.04], 0.05 [0.02, 0.07], and 0.09 [0.07, 0.12] for the 1946, 1958, and 1970 birth cohorts, respectively. Increases in distress were larger among women than men, widening the pre-existing inequalities observed in the pre-pandemic peak and in the most recent pre-pandemic assessment.

**Conclusions and Relevance:** Pre-existing long-term psychological distress trajectories of adults born between 1946 and 1970 were disrupted during the COVID-19 pandemic, particularly among women, who reached the highest levels ever recorded in up to 40 years of follow-up data. This may impact future trends of morbidity, disability, and mortality due to common mental health problems.

## Introduction

Mental disorders are among the leading global contributors to years lived with disability.^1,2^ Growing evidence suggests that this may have worsened given the impact of the COVID-19 pandemic and the restriction measures put in place to control its spread, on mental health, including depression, anxiety, and, more generally, psychological distress.^3-7^

In the UK, results from 11 longitudinal population-based studies show that psychological distress levels have been, overall, higher throughout the first year after the pandemic onset compared to pre-pandemic levels.^8^ This complements earlier evidence focused on the initial stages of the pandemic, where worsening levels of mental health outcomes –particularly anxiety and distress levels– were reported.^9-13^ Although these studies are crucial to understand whether population mental health has worsened during the pandemic, they do not provide evidence on where these changes stand in relation to pre-existing long-term mental health trajectories.

Evidence prior to the pandemic using data from the British birth cohorts has shown that, throughout adulthood, there seems to exist an upwards trend in the long-term psychological distress trajectories by middle age (age 30-45), and a decrease towards older age.^14,15^ By extending these analyses to include data collected during the first year after the COVID-19 pandemic onset, we aims to understand whether the changes in distress reflect a continuation or an alteration/disruption of these pre-pandemic trends, which may have implications for future trends of morbidity, disability, and mortality.^2,16^

Moreover, evidence on the changes in mental health outcomes suggest that women and younger adults have been generally hit harder by the pandemic,^9-13^ in agreement with global evidence.^17^ By analysing these long-term psychological distress trajectories across generations and sexes, we also aim to investigate whether there are inequalities in the potential disruption of the pre-existing long-term trends across generations and sexes.

## Methods

### Sample and procedure

We used data from three British birth cohorts: the National Survey of Health and Development (NSHD),^18^ the National Child Development Study (NCDS),^19^ and the British Cohort Study (BCS70),^20^ representing people born in a single week in Britain in 1946, 1958, and 1970, respectively. Life-course data from the studies (**eAppendix 1**) were augmented with the COVID-19 Survey,^21^ which collected relevant information regarding the pandemic on the members of these cohort studies at three time-points: May 2020 (during the first national lockdown), September-October 2020 (between the first and second national lockdowns), and February-March 2021 (during the third national lockdown). NCDS data was further augmented with data on 1,366 participants from age 62 sweep fieldwork, which started in January 2020 and had to be paused due to the pandemic onset.^22^ In this study, we focused on cohort members who took part in the COVID-19 Survey in at least one time-point. Ethical approval was obtained from the National Health Service (NHS) Research Ethics Committee, and all participants provided informed consent.

### Measures

#### Psychological distress

In both NCDS and BCS70, psychological distress was measured with a nine-item version of the Malaise Inventory ^23,24^ at all time-points, including the COVID-19 survey. Previous studies have shown that, up to the most recent pre-pandemic assessment in these two cohorts, these nine items reflected equivalently the same construct over time and across cohorts and sexes.^15,25^ In NSHD, different questionnaires were used over time, both prior to and during the COVID-19 pandemic. The Present State Examination (PSE)^26^ was used at age 36; the Psychiatric Symptoms Frequency (PSF, based on the PSE)^27^ at age 43; and, from then onwards, two different versions of the General Health Questionnaire: the GHQ-28 at ages 50-69, and the GHQ-12 during the COVID-19 Survey, corresponding to ages 74-75.^28^ The item harmonisation procedure reported elsewhere^14,29^ was implemented where items from these different questionnaires were mapped to specific distressing experiences. The two-item versions of the Patient Health Questionnaire (PHQ-2)^30^ and the Generalized Anxiety Disorder (GAD-2)^31^ questionnaires were administered during the COVID-19 survey in all cohorts in addition to their corresponding psychological distress measures. Additional information on the measures and on the harmonisation process used is available in **eAppendix 1** and **eAppendix 2**, respectively.

Due to the wide range of different measures of psychological distress across cohorts (NSHD vs NCDS and BCS70) and within NSHD, we operationalised psychological distress as a factor score (continuous). This included all cohorts and leveraged the existence of a common set of indicators of psychological distress (PHQ-2 and GAD-2) across the three cohorts during the COVID-19 Survey waves, in addition to the cohort-specific items. The common items were used as ‘anchor items’ to estimate a psychological distress factor and derive the corresponding factor scores across cohorts and time-points using an Item Response Theory (IRT) based linking approach.^32^ As sensitivity checks, we used additional psychological distress operationalisations (**eAppendix 3**).

Information on the cohort members’ biological sex as recorded at birth was used.

### Data analyses

#### Measurement invariance/equivalence testing

To ensure that changes in the psychological distress levels were not due to changes in the properties of the measurement tools over time and across cohorts and sexes, a measurement invariance/equivalence testing procedure was implemented using a Structural Equation Modelling (SEM) framework.^33^ Further details on the measurement invariance testing procedure used, along with its results, are available in **eAppendix 4**.

#### Derivation of factor scores

After obtaining evidence on the invariant measurement properties of the four identical psychological distress indicators in the COVID-19 Survey waves (the GAD-2 and PHQ-2 items) (**eAppendix 4**), these four indicators were pooled, along with the cohort-specific psychological distress indicators (**eAppendix 5**). A Full Information Maximum Likelihood (FIML) estimation, corrected for the clustering induced by the longitudinal design (MLR), was used. This enabled factor scores for each time-point with at least partial information available to be obtained.^34^ The same procedure was implemented in the additional sensitivity checks within NSHD, where the seven previously harmonised symptoms^14,29^ were used as indicators of a latent psychological distress factor, and factor scores were derived for all time-points with at least partial information, including the COVID-19 Survey waves where four out of the seven previously harmonised symptoms were missing by design.

#### Trajectories of psychological distress

We used a multilevel growth curve modelling approach to analyse the trajectories of psychological distress under the different outcome operationalisations. Unadjusted models were estimated separately for each cohort. The models were also estimated including an interaction term between each growth parameter and birth sex, to account for inequalities in these trajectories within cohorts in line with the abovementioned evidence. The random part of all these models included the variation in the initial levels (random intercepts) but not in the change over time (random slopes) as the inclusion of this additional random effect led to convergence issues. To answer the counterfactual question of what the distress levels would have been had the COVID-19 pandemic not occurred, models estimated with data only up to the most recent pre-pandemic assessment (2015, early 2020, and 2016 in NSHD, NCDS, and BCS70, respectively) were used to obtain projections of the distress levels in 2020 and 2021. Further details on the analytical approach are available in **eAppendix 6**.

We obtained the standardised mean differences (SMD) in the factor scores between the peak during the pandemic and two relevant pre-pandemic time-points: the pre-pandemic peak by midlife^14^ and the most recent pre-pandemic assessments. These SMDs were obtained for the three cohorts both overall and by birth sex. We then used a difference-in-differences (DiD) approach to explore whether the sex differences had changed at the pandemic peak compared to those pre-pandemic points (pre-pandemic peak and most recent pre-pandemic assessment).

To account for the differential probability of participating in the COVID Survey waves, and thus restore sample representativeness to the target population, all models were estimated using an inverse probability weighting (IPW) approach. The weights were generated for each of the three COVID Survey waves based on personal characteristics and the history of previous participation.^35^ In NSHD, these weights were combined with the corresponding design weights.^18^ Additional information on the derivation of these weights and their effectiveness to restore sample representativeness is available in the COVID-19 Survey User Guide.^21^

SEM models (measurement models to test invariance/equivalence and to obtain factor scores) were estimated in Mplus version 8.6.^36^ Multilevel growth curve models were estimated in Stata MP version 17.0.^37^

## Results

After excluding participants who did not take part in any of the COVID-19 survey waves, the overall sample comprised *N*=16,389 participants from NSHD (*n*=2,175, 52.8% women), NCDS (*n*=7,446, 52.4% women), and BCS70 (*n*=6,768, 56.2% women) (**eAppendix 7**). Number of repeated observations ranged from 1 to 8 in NSHD (median=7), NCDS (median=6), and BCS70 (median*=*6).

### Trajectories of distress as factor scores

A clear period effect was observed in all three cohorts during the COVID-19 pandemic, which indicated a disruption to the psychological distress trajectories that had been observed prior to the start of the pandemic across the cohorts. The unadjusted marginal predicted mean psychological distress levels (**Figure 1**) increased from the pandemic onset onwards and, by September/October 2020 (between first and second national lockdowns, second of the last three points in the figure), they had reached (NSHD) or exceeded (NCDS and BCS70) the highest average distress levels in the pre-pandemic trajectories. A decrease was then observed towards the last point, corresponding to February/March 2021 (during third national lockdown) in both NSHD and BCS70, whereas mean levels slightly increased further in NCDS. In all cases, distress levels by the last observation were notably higher than the last pre-pandemic levels. Models’ coefficients and marginal predicted levels using the cross-cohort factor score operationalisation are available in **eAppendix 8**.

**Figure 1.**
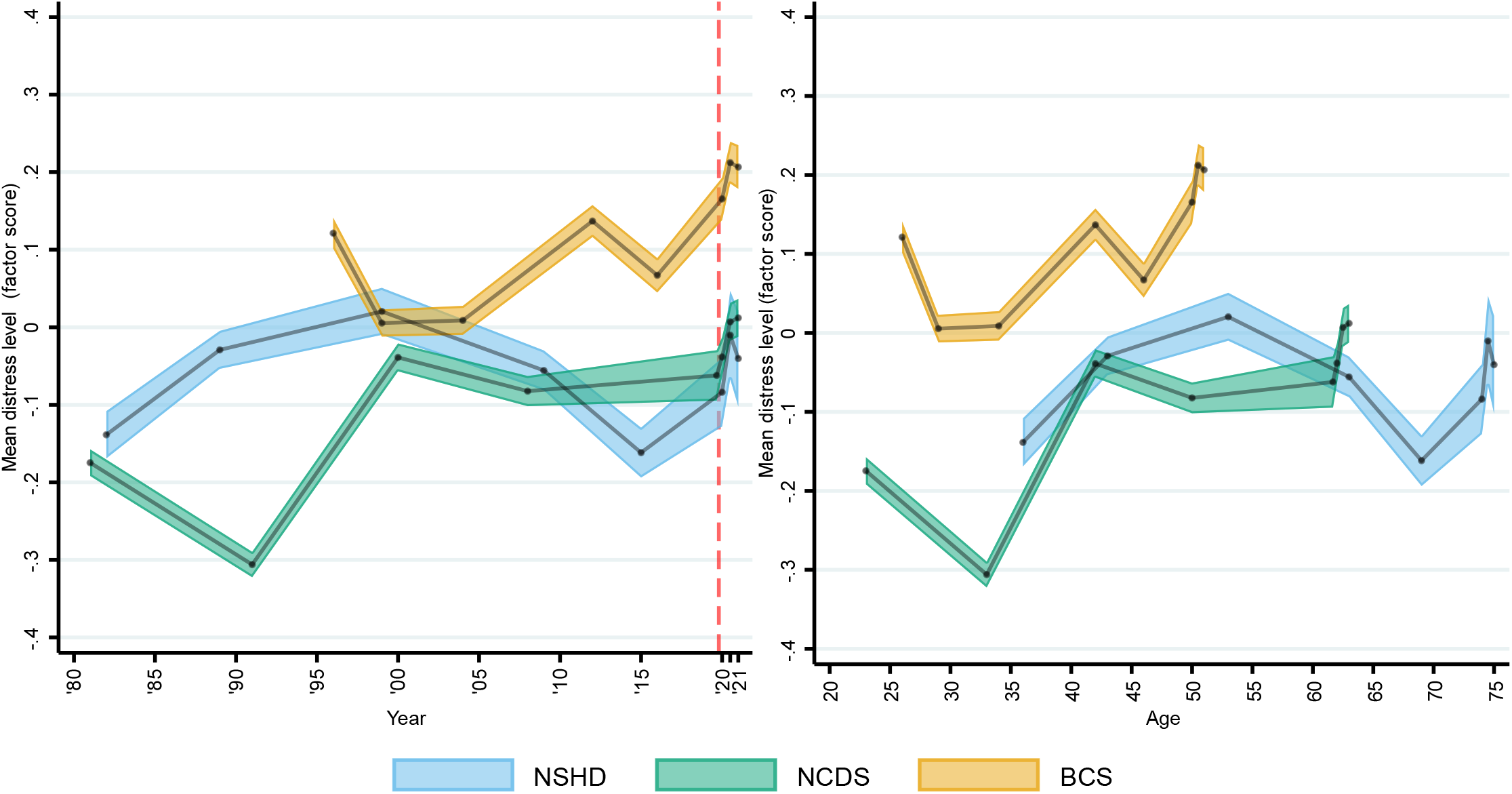
Marginal mean psychological distress cross-cohort factor scores over time (year and age). Note: Unadjusted results. 95% confidence intervals are indicated in lighter shaded areasBCS70: 1970 British Cohort Study NCDS: 1958 National Child and Development Study; NSHD: 1946 National Survey of Health and Development. The dashed line represents the first nationwide lockdown enforced in March 2020.

The psychological distress projections obtained from the models using only pre-pandemic data (**Figure 2**) also supported the notion of an alteration in the long-term trajectories of distress with the pandemic onset.

**Figure 2.**
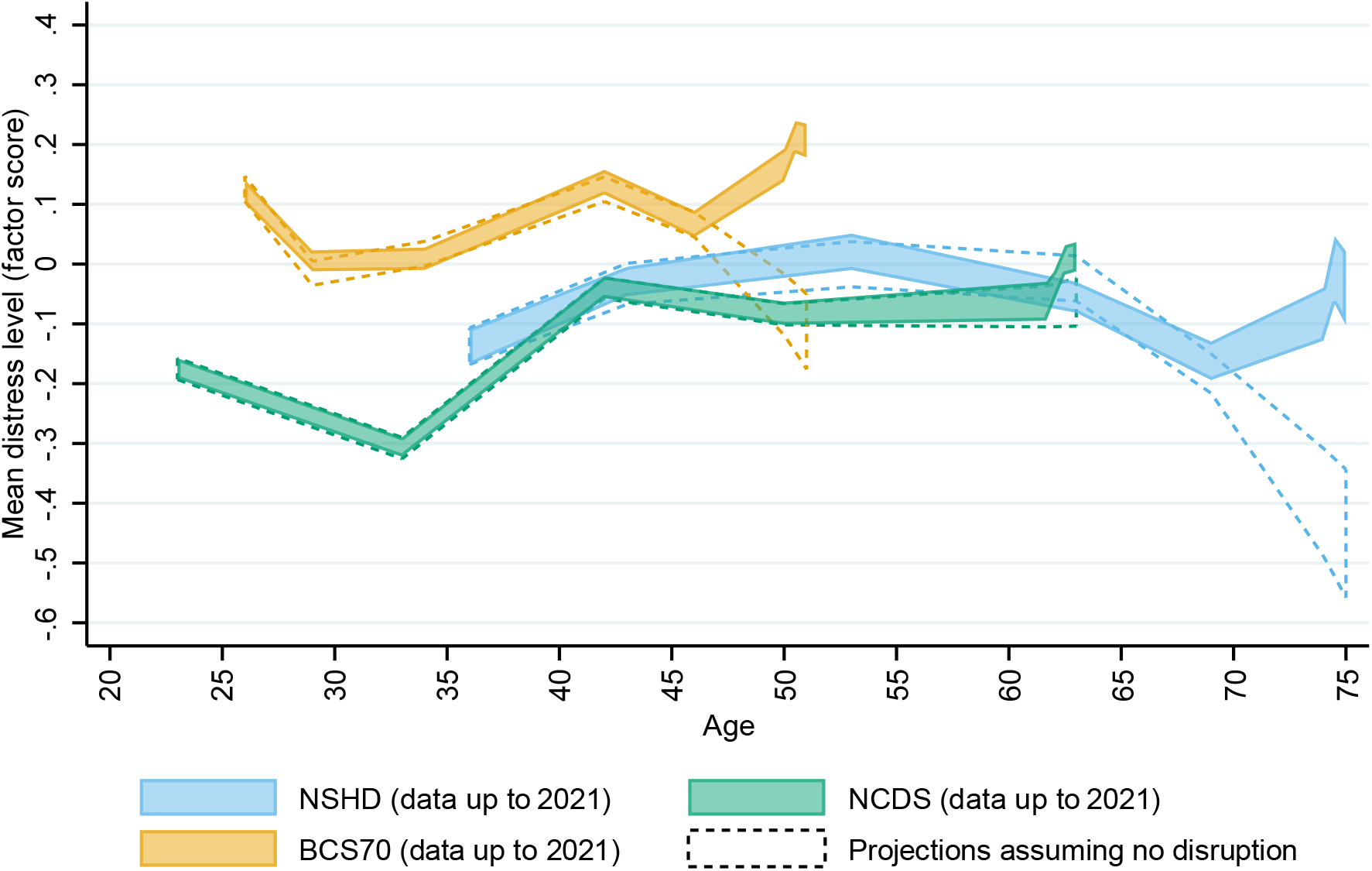
Projections of mean number of psychological distress symptoms from pre-pandemic data (assuming no disruption) and marginal predicted levels from the analyses using data collected during the pandemic. Note: Unadjusted results. All areas correspond to 95% confidence intervals. BCS70: 1970 British Cohort Study; NCDS: 1958 National Child and Development Study; NSHD: 1946 National Survey of Health and Development. Projections assuming no disruption are based on data up to2015 (NSHD), 2020 (NCDS), and 2016 (BCS70).

The interaction terms between birth sex and the parameters corresponding to the changes during the pandemic (spline 2, **eTable 8.1**) were only statistically significant for NCDS (B_NCDS,spline2linear*women_=0.70 [0.32, 1.08], *p*<0.001; B_NCDS,spline2quadratic*women_=-0.87 [-1.55, -0.20], *p*=0.011; B_NCDS,spline2cubic*women_=0.33 [0.01, 0.65], *p*=0.043), evidencing a significantly different trajectory during the pandemic between men and women. The visual exploration of the marginal predicted levels by birth sex obtained from these models (**eFigure 8.1**) confirmed this, showing differences in the trajectories during the pandemic across the other two cohorts as well.

Figure 3 shows the SMD in the distress factor scores between September/October 2020 and the pre-pandemic peak in midlife (left section) and the most recent pre-pandemic assessment (right section), both overall and by birth sex. Overall, SMD were larger when compared to the most recent pre-pandemic assessment (SMD_NSHD,recent_=0.17 [0.06, 0.28], *p*<0.001; SMD_NCDS,recent_=0.11 [0.07, 0.16], *p=*0.003; SMD_BCS70,recent_=0.11 [0.05, 0.16], *p*<0.001) than to the pre-pandemic peak in midlife (SMD_NSHD,pre-peak_=-0.02 [-0.07, 0.04], *p*=0.518; SMD_NCDS,pre-peak_=0.05 [0.02, 0.07], *p*<0.001; SMD_BCS70,pre-peak_=0.09 [0.07, 0.12], *p*<0.001), and differences with the pre-pandemic peak in midlife were larger in younger cohorts. In all cases, the overall SMD concealed the underlying sex inequalities, with women showing larger differences than men. The DiD analysis supported this observation, showing that, in all cohorts, sex inequalities had widened by September/October 2020 compared to those observed in the pre-pandemic peak in midlife (DiD_NSHD,sex,pre-peak_=0.17 [0.06, 0.28], *p*=0.002; DiD_NCDS,sex,pre-peak_=0.11 [0.07, 0.16], *p*<0.001; DiD_BCS70,sex,pre-peak_=0.11 [0.05, 0.16], *p*<0.001) and in the most recent pre-pandemic assessment (DiD_NSHD,sex,recent_=0.14 [0.04, 0.24], *p*=0.005; DiD_NCDS,sex,recent_=0.15 [0.08, 0.23], *p*<0.001; DiD_BCS70,sex,recent_=0.09 [0.05, 0.14], *p*<0.001).

**Figure 3.**
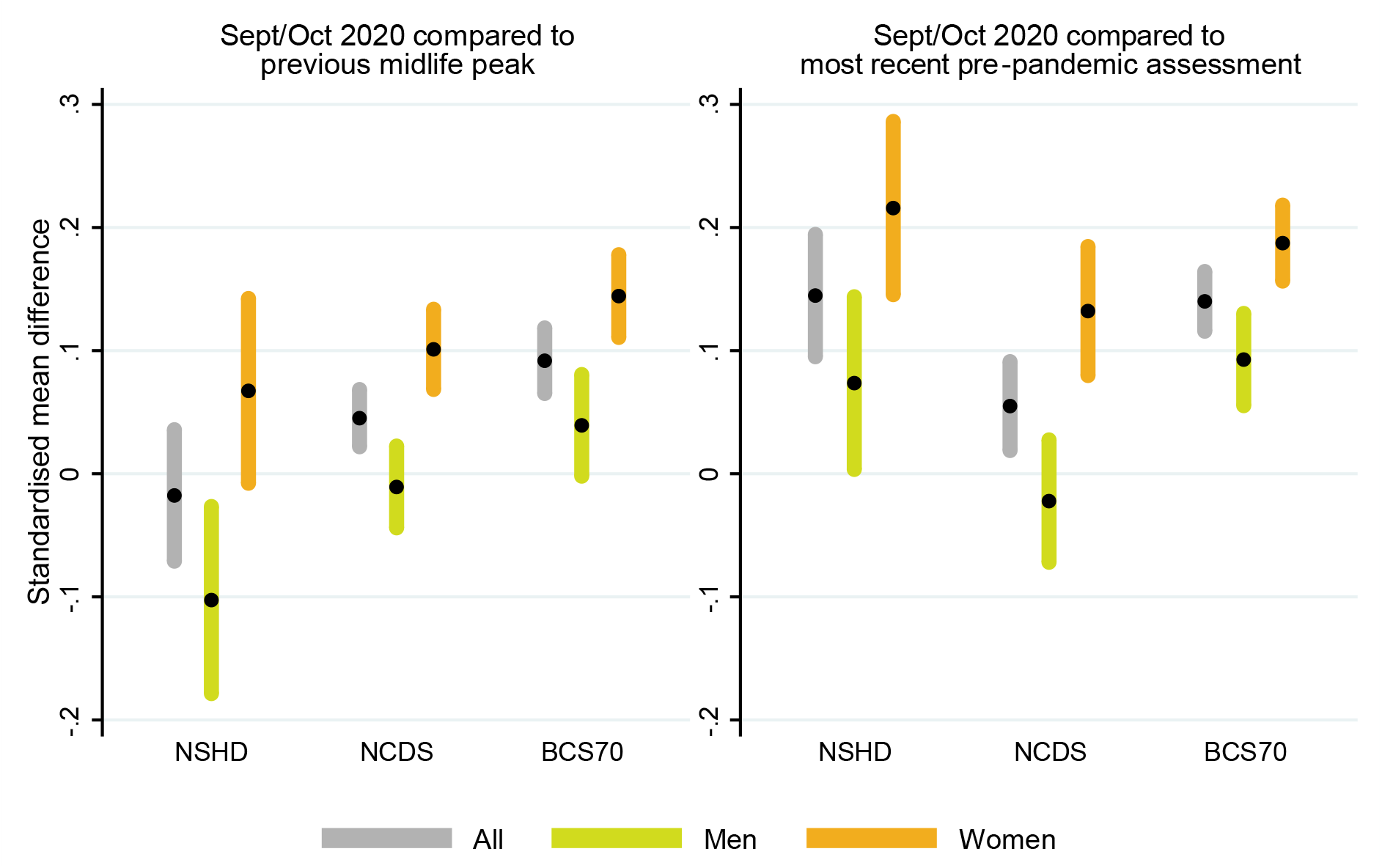
Standardised mean difference in cross-cohort factor scores between September/October 2020 and pre-pandemic peak in midlife and most recent pre-pandemic assessment. Note: Unadjusted results. 95% confidence intervals are indicated in shaded areasBCS70: 1970 British Cohort Study; NCDS: 1958 National Child and Development Study; NSHD: 1946 National Survey of Health and Development. Previous midlife peaks correspond to years1999 (NSHD), 2000 (NCDS), and 2012 (BCS70). Most recent pre-pandemic assessments correspond to2015 (NSHD), January/March 2020 (NCDS), and 2016 (BCS70).

### Sensitivity checks

Analyses performed with the observed ‘number of symptoms’ operationalisation (**eAppendix 9**), the ‘caseness’ operationalisation (**eAppendix 10**), and the factor scores derived from the seven harmonised indicators within NSHD (**eAppendix 11**) provided very similar results as those found in the main analyses. In all these alternative operationalisations, psychological distress levels in all cohorts reached an all-time peak by September/October 2020, and a larger alteration with the pandemic onset was observed in the oldest cohort (NSHD) when using the ‘caseness’ operationalisation.

## Discussion

Our study aimed to investigate if there had been a disruption in the pre-existing long term psychological distress trajectories of the UK adult population during the COVID-19 pandemic, and to analyse if such disruptions were related to the pandemic. We used a triangulation approach in the three oldest British birth cohorts, born in 1946, 1956 and 1970, using observed data on different distress operationalisations before and during the pandemic, obtaining projections based on pre-pandemic data, and examining the differences between relevant time-points before and after the pandemic onset. All these different approaches suggest that the pre-existing long-term distress trajectories, which had reached their peak by midlife (around age 40-50), were altered during the first year of the COVID-19 pandemic. Distress levels increased with respect to pre-pandemic levels, in most cases reaching the highest average levels over the life-course by September/October 2020. Although average distress levels tended to decrease afterwards, they were notably higher than before the pandemic onset one year after the first national lockdown. Our study also suggests that this pattern was significantly worse in women than in men regardless of age. The emergence of a new peak in the distress trajectories may increase the morbidity, disability, and mortality due to common mental health problems, which were already among the leading causes of global burden of disease without accounting for this new peak,^1,2,16^ with women likely being disproportionately affected by these potential increases, which may result in even greater inequalities by sex.

The finding of an increase in psychological distress with regard to pre-pandemic levels is consistent with previous evidence showing an overall deterioration in mental health outcomes in the UK adult population,^10-12^ or in adults over the age of 50.^9,13^ The difference between the levels reached during the pandemic and the corresponding pre-pandemic peak was generally larger among younger cohorts regardless of sex. Considering that younger cohorts had higher levels of distress throughout the adulthood before the pandemic,^14,15^ these results may also point at future increasing inequalities by generation. However, this finding was not consistent across the additional psychological distress operationalisations in this study. This, along with the steady levels by the last time-point in NCDS, compared to the decreasing levels observed in the other two cohorts, points at the need for further monitoring and study of these generational inequalities.

In line with previous evidence,^9-13,17^ we found that women had worse distress levels than men throughout the COVID-19 pandemic, as noted. Although distress levels were already higher in women throughout adulthood, the observed period effect was larger in women. By September/October 2020, women’s distress levels exceeded the levels observed in the most recent pre-pandemic assessment in all cohorts, and exceeded (or reached, whilst men did not) the levels observed in the pre-pandemic peak. Our study suggests that sex inequalities in psychological distress during the pandemic may not just be a continuation of pre-pandemic long-term inequalities, suggesting that these widened during the pandemic. Women have taken a disproportionately larger share of the unpaid care work responsibilities arising from pandemic control measures, including housework, home-schooling, and caring responsibilities.^38,39^ Rates of domestic and gender-based violence and abuse have also reportedly increased during lockdowns.^40,41^ Moreover, recent evidence suggests that, in addition to first-hand bereavements through the loss of loved ones during the pandemic, the mental health of women aged 50 and older may have also been affected by the collective, larger-scale death toll of the pandemic,^42^ which in the UK remains one of the highest in Europe.^43^ These different factors may partly explain the larger disruption of the pre-existing long-term distress trajectories experienced by women during the pandemic.

### Strengths and limitations

Our study has several strengths. It is, to the best of our knowledge, the longest longitudinal study of psychological distress trajectories to date, following the same individuals for up to 40 years and showing the unique effect of the pandemic over the life-course. Using data from birth cohorts enabled us to understand the potential impact of the COVID-19 pandemic in the context of the distress levels experienced by the same individuals throughout their adulthood prior to the pandemic’s onset, with data collected prospectively, and a high degree of generalisability, due to the cohorts being nationally representative. Through the use of an IRT-based linking approach leveraging the existence of common distress indicators across the birth cohorts used, we were able to increase the comparability across these cohorts compared to previous evidence.^14^ By using multiple operationalisations of psychological distress, including but not limited to binary outcomes, we qualify previous evidence focused on the latter,^8^ showing that our main results are robust to these different operationalisations while acknowledging the differences across them. Our study also has limitations. As expected in cohort designs, our study suffered from high proportions of attrition with respect to the original samples. To limit the impact of attrition, we used non-response weights which have been found to be effective at restoring sample representativeness with respect to the characteristics of the respective target populations: those born in the UK in 1946, 1958, or 1970, alive and residing in the UK.^21^ However, although this study’s results may be representative of these target populations, they may not be generalisable to other sections within the UK adult population (such as migrants and ethnic minority groups, which by 2019 made up about 14% of the UK’s population^44^ and 15% of the population in England and Wales,^45^ respectively) and countries different than the UK (particularly those with different cultural, socioeconomic, and political characteristics).^46^ Finally, it was obviously not possible to include a contemporaneous control group unexposed to the pandemic in the analysis. Although we used projections based exclusively on pre-pandemic data in order to resemble the expected distress levels had the pandemic not occurred, we are aware that these counterfactual analyses have their own limitations: first, they are based on a small number of pre-pandemic data points, which limited the granularity of the predictions; second, the last time-point used in NCDS corresponded to the period just before the national lockdown came into force, and therefore participants may have already been preoccupied with the pandemic. This may partly explain why these projections showed a substantially smaller increase in NCDS, but further research is needed to clarify whether this was the case. It is also possible that the observed period effect was the result of pre-existing trends and unrelated to the pandemic. However, this is unlikely considering the triangulation of the results from the different analyses using data from three different cohorts, which support the notion of a pandemic-related disruption to long-term psychological distress trajectories.

## Conclusions

This longitudinal study conducted with three prospective UK birth cohorts shows that pre-existing long-term psychological distress trajectories of adults born between 1946 and 1970 were disrupted during the COVID-19 pandemic, reaching or exceeding the highest levels previously recorded in up to 40 years of follow-up data. This disruption may lead to increases in the morbidity, disability, and mortality due to common mental health problems, particularly among women, whose distress trajectories have been disproportionately altered, resulting in growing sex inequalities. Public policies aimed at the provision of support and continued monitoring of population mental health are crucial in light of these results, with a focus on those most disproportionately impacted.

## Data Availability

Data for the British Cohort Study (BCS70, SN 8547), the National Child Development Study (NCDS, SN 6137), and the COVID-19 surveys (SN 8658) are available from the UK Data Service (https://ukdataservice.ac.uk/). Deidentified data and documentation on the National Survey of Health and Development (NSHD) are available under request (https://www.nshd.mrc.ac.uk/data).

https://www.nshd.mrc.ac.uk/data

## Authors’ contributions

DM, JD, and GBP conceived the study. DM conducted the data curation, methodology, analysis, and visualisation, and wrote the original draft. JD and GBP supervised the project. All authors reviewed, edited, and approved the final manuscript.

## Author access to data

DM had full access to all the data in the study and takes responsibility for the integrity of the data and the accuracy of the data analysis.

## Conflicts of interest

None to declare.

## Sources of funding and support

This paper represents independent research part supported by the Economic and Social Research Council (ESRC) Centre for Society and Mental Health at King’s College London [ES/S012567/1]. DM, HLF, SLH, CM, JD, and GBP are part supported by the ESRC Centre for Society and Mental Health at King’s College London [ES/S012567/1]. SLH, CM, and JD are also supported by the National Institute for Health Research (NIHR) Biomedical Research Centre at South London and Maudsley NHS Foundation Trust and King’s College London and JD is also supported by the NIHR Applied Research Collaboration South London (NIHR ARC South London) at King’s College Hospital NHS Foundation Trust. MR is supported by the Medical Research Council (MRC) [MC_UU_00019/3]. The views expressed are those of the authors and not necessarily those of the ESRC, NIHR, the Department of Health and Social Care, MRC, or King’s College London.

The British Cohort Study 1970 and National Child Development Study 1958 are supported by the Centre for Longitudinal Studies, Resource Centre 2015-20 grant [ES/M001660/1] and a host of other co-funders. The NSHD cohort is hosted by the MRC Unit for Lifelong Health and Ageing at UCL funded by the MRC [MC_UU_00019/1Theme 1: Cohorts and Data Collection]. The COVID-19 data collections in these five cohorts were funded by the UKRI grant Understanding the economic, social and health impacts of COVID-19 using lifetime data: evidence from 5 nationally representative UK cohorts [ES/V012789/1].

## Role of funder(s)/sponsor(s)

The funders had no role on the design and conduct of the study; collection, management, analysis, and interpretation of the data; preparation, review, or approval of the manuscript; or decision to submit the manuscript for publication.

## Other acknowledgements

We would like to thank all individuals who participated in the three birth cohort studies for so generously giving up their time over so many years, and all the study team members for their tremendous efforts in collecting and managing the data. The authors would also like to thank Dr Dawid Gondek for providing very helpful code for some of the data management and statistical analyses used in this study.

## Notes

### Competing Interest Statement

The authors have declared no competing interest.

### Author Declarations

The most recent sweeps of the British Cohort Study (BCS70), the National Child Development Study (NCDS), and the National Survey of Health and Development (NSHD) have all been granted ethical approval by the National Health Service (NHS) Research Ethics Committee and all participants have given informed consent. No additional ethical approval was necessary for this secondary data analysis.

