## Supplementary Material for "Disruption of long-term psychological distress trajectories during the COVID-19 pandemic: evidence from three British birth cohorts"

**eAppendix 1. Information on the psychological distress measures used in the study**

| Cohort | Measure | Details | 'Caseness' threshold | Psychometric and clinical properties |
|---|---|---|---|---|
| **NSHD** | Present State Examination (PSE) | Nurse-administered clinical examination assessing the frequency and severity of a range of psychiatric symptoms in the preceding month. Used at age 36. | 5 or higher on the Index of Definition.[1] | Tested in general population: high agreement with clinical diagnosis of common mental health disorders (~90%); high concurrent validity with other measures of psychological distress.[1] |
| | Psychiatric Symptoms Frequency (PSF) | Nurse-administered questionnaire (18 items) assessing symptoms of anxiety and depression during the preceding year. 5-point scale from 0 ("Never in the last year") to 5 ("Every day in the last year"). Used at age 43. | 23 or higher on the summed up score.[2] | Tested in general health care/general population: Cronbach's alpha = 0.88; all items identified a common factor; AUC against reports of contact with doctor/use of prescribed medication for 'nervous or emotional trouble or depression' = 0.84-0.86.[2] |
| | General Health Questionnaire, 28-item version (GHQ-28) | Self-administered questionnaire (28 items) assessing symptoms of anxiety in depression in the preceding 4 weeks. 4-point scale recoded into binary values.[3] Used at ages 50-69. | 6 or higher score.[3] | Tested in general health care (adults): Cronbach's alpha = 0.82-0.86; AUC against diagnosed psychiatric morbidity = 0.88.[3] |
| | General Health Questionnaire, 12-item version (GHQ-12) | Self-administered questionnaire (12 items) assessing whether symptoms of anxiety and depression were 'recently' experienced. 4-point scale recoded into binary values.[3] Used at ages 74-75. | 2 or higher score.[3] | Tested in general health care (adults): AUC against diagnosed psychiatric morbidity = 0.88.[3] |
| **NCDS and BCS70** | Malaise Inventory | Self-administered questionnaire (9 items) assessing whether distressful experiences are 'generally' experienced. Used at all ages from 23 (NCDS) and 26 (BCS70) onwards. | 4 or higher on the summed-up score | Tested in general population, the full 24-item version showed an AUC against self-reported diagnosed psychiatric morbidity = 0.77-0.79.[4] The 9-item version correlates highly (r=0.91-0.92) with the 24-item version.[5] |
| **NSHD, NCDS, and BCS70** | Generalized Anxiety Disorder, 2-item | Self-administered questionnaires (2 items) assessing two core anxiety and depressive | Not applicable. These items are used in the factor score approach with the three cohorts as indicators | Tested in general health care (adults): AUC against major depressive disorder diagnostic = 0.93, against |

| | | | | |
|---|---|---|---|---|
| | version (GAD-2) | symptoms, respectively, over the last 2 weeks. 4-point scale from 1 (“Not at all”) to 4 (“Nearly every day”). Used at all three COVID-19 Survey waves. | of the latent level of psychological distress due to their presence in all three cohorts at the same three time points (‘anchor’ items). Therefore, no ‘caseness’ thresholds are used. | any depressive disorder = 0.90; correlation with functional impairment due to mental health issues (r=0.70).[6] |
| | Patient Health Questionnaire, 2-item version (PHQ-2) | | | Tested in general health care (adults): AUC against generalised anxiety disorder = 0.91, against any anxiety disorder = 0.85.[7] |

*Note*. Adapted from Gondek at al.[8]

**eAppendix 2. Harmonised items across psychological distress measures in NSHD**

| **Symptom** | **Present State Examination (PSE)** | **Psychiatric Symptom Frequency (PSF)** | **General Health Questionnaire, 28-item version (GHQ-28)** | **General Health Questionnaire, 12-item version (GHQ-12)** |
|---|---|---|---|---|
| Low Mood | Do you keep reasonably cheerful or have you been very depressed or low spirited recently? (rate depressed mood) | Over the last year have you been in low spirits or felt miserable? | Have you recently been able to enjoy your normal day-to-day activities? | Have you recently been feeling unhappy or depressed? * |
| Fatigue | Have you been exhausted and worn out during the day or evening even when you haven't been working very hard? (rate tiredness/exhaustion) | Over the last year have there been days when you tired out very easily? | Have you recently been feeling in need of a good tonic? | |
| Tension | Do you often feel on edge, keyed up, mentally tense or strained? (rate nervous tension) | Over the last year have you felt on edge, keyed up or mentally tense? | Have you recently felt constantly under strain? | Have you recently felt constantly under strain? |
| Panic | Have you had times when you felt shaky or you heart pounded or you felt sweaty and you simply had to do something about it? | Over the last year have you been in situations when you felt shaky or sweaty or your heart pounded or you could not get your breath? | Have you recently been getting scared or panicky for no good reason? | |
| Hopelessness | How do you see the future? (rate hopelessness) | Over the last year have you had the feeling that the future does not hold much for you? | Have you recently felt that life is entirely hopeless? | |
| Health anxiety | Do you tend to worry over your physical health? (rate hypochondriasis) | Over the last year have you been frightened or worried about becoming ill or about dying? | Have you recently felt that you are ill? | |
| Sleep problems | Have you had any trouble getting off to sleep in the last month?<br>(rate delayed sleep) | Over the last year have you had trouble getting off to sleep? | Have you recently lost much sleep over worry? | Have you recently lost much sleep over worry? |
| *Response options* | Symptom not present/ Symptom definitely present during past month, but of moderate clinical intensity/ Intense form of symptom present for more than 50% of past month | Never/ Occasionally/ Sometimes/ Quite often/ Very often/ Always | Not at all/No more than usual/ Rather more than usual/ Much more than usual | Not at all/No more than usual/ Rather more than usual/ Much more than usual |

*Note*. Adapted from Gondek et al.[8] and McElroy et al.[9] Additional details on the harmonisation procedure are available in McElroy et al.[9] * Although the question used to represent the "low mood" symptom was present in both the GHQ-28 and GHQ-12, this item was deemed inadequate to adequately reflect the person's mood at the first COVID-19 Survey wave, seemingly being more reflective of the current enforced restrictions legally limiting the ability of the person to perform their normal day-to-day activities. More information on this is available in the eAppendix 3 in this same document.

**eAppendix 3. Additional psychological distress operationalisations used in sensitivity checks**

As sensitivity checks, we used alternative psychological distress operationalisations, in addition to the main operationalisation as a factor score. First, we operationalised psychological distress as the number of symptoms present (discrete) at each time-point. This could be directly done in NCDS and BCS70 due to the use of the same instrument across cohorts and over time; and relied on three out of the seven previously harmonised symptoms that were present across all data collection points in NSHD due to the change in the version of the GHQ used in the COVID-19 Survey. Thus, the potential number of symptoms ranged from 0 to 9 in NCDS and BCS70, and from 0 to 3 in NSHD. Second, psychological distress was operationalised as 'caseness' (binary), using each of the measurement tools' recommended thresholds (**eAppendix 1** in this document). Finally, an additional factor approach was implemented in NSHD using the seven previously harmonised symptoms as indicators of a latent psychological distress factor.

Trajectories were modelled using different multilevel growth curve models depending on the nature of the outcome: linear models for the factor scores operationalisations (continuous), Poisson models for the number of symptoms operationalisation (discrete), and logistic models for the 'caseness' operationalisation (binary)

Analyses using NSHD data accounted for likely reporting bias during the COVID-19 survey of one of the GHQ-12 items (item 7 from GHQ-12: "Have you recently been able to enjoy your normal day to day activities"), which seemed to be more reflective of the lockdown restrictions in place than of the person's distress level (see below). This item was therefore replaced by a different item on low mood (item 9: "Have you recently been feeling unhappy or depressed?") in the approaches using harmonised symptoms.

The initial exploration of the proportion of endorsement of each of the GHQ-12 items, after recoding the responses following the GHQ-scoring system,[3] showed an unusually high endorsement (51.7%) of item 7 in the COVID-19 Survey wave 1 (May 2020). This item corresponds to the question "Have you recently been able to enjoy your normal day to day activities?", which was objectively impaired considering the ongoing nationwide lockdown at the assessment time point. The difference can be clearly observed in **eFigure 3.1**, which includes all the GHQ-12 items, with those marked with an asterisk (*) having been reversely coded to reflect psychological distress.

Since item 7 may be more reflective of the external conditions than of the psychological distress level of the individual, as most people could not legally perform their normal day to day activities back then, we were cautious when including it in the different operationalisations of psychological distress.

In the 'caseness' (binary) approach, this item may have a strong impact on the proportion of 'cases' using the recommended threshold of 2+ endorsed items, thus potentially leading to overestimation of those proportions. We compared the 'caseness' prevalence estimates resulting from three different operationalisations: a) usual approach, keeping the problematic item in and the threshold at 2+; b) dropping the item and keeping the threshold at 2+; and c) dropping the item and lowering the threshold to 1+. The weighted proportion estimates of 'caseness' at each of the three COVID-19 Survey waves is shown in **eTable 3.1**.

Operationalisation A (usual approach) showed a very high prevalence estimate during the first survey wave, an increase that may be mostly driven by item 7 as the comparison with the other two operationalisations, which follow the inverted U-pattern, suggest. Operationalisation B showed that inverted U-pattern, consistent with what has been found in the alternative operationalisations and cohorts, and showed relatively similar results to the usual approach (operationalisation A) at survey waves 2 and 3. Operationalisation C led to the highest prevalence estimates of all operationalisations, despite showing the expected inverted U-pattern. Thus, operationalisation B was selected as the optimal approach to 'caseness' in NSHD during the COVID-19 Survey waves.

In the 'number of symptoms present' (discrete) and the factor score (continuous) approaches, which were based on the harmonisation of items mapped to specific distressful experiences, the problematic item (item 7) had been previously used as the indicator of choice for low mood,[8,9] present in the 28-item version of the GHQ used in that study. An alternative item (item 9: "Have you recently been feeling unhappy or depressed?") could be mapped to low mood in the 12-item version and was therefore used as the indicator of choice in the three COVID-19 Survey assessments (see **eAppendix 2** in this document).

**eFigure 3.1. Proportion of NSHD participants endorsing each GHQ-12 item in the COVID-19 Survey waves**

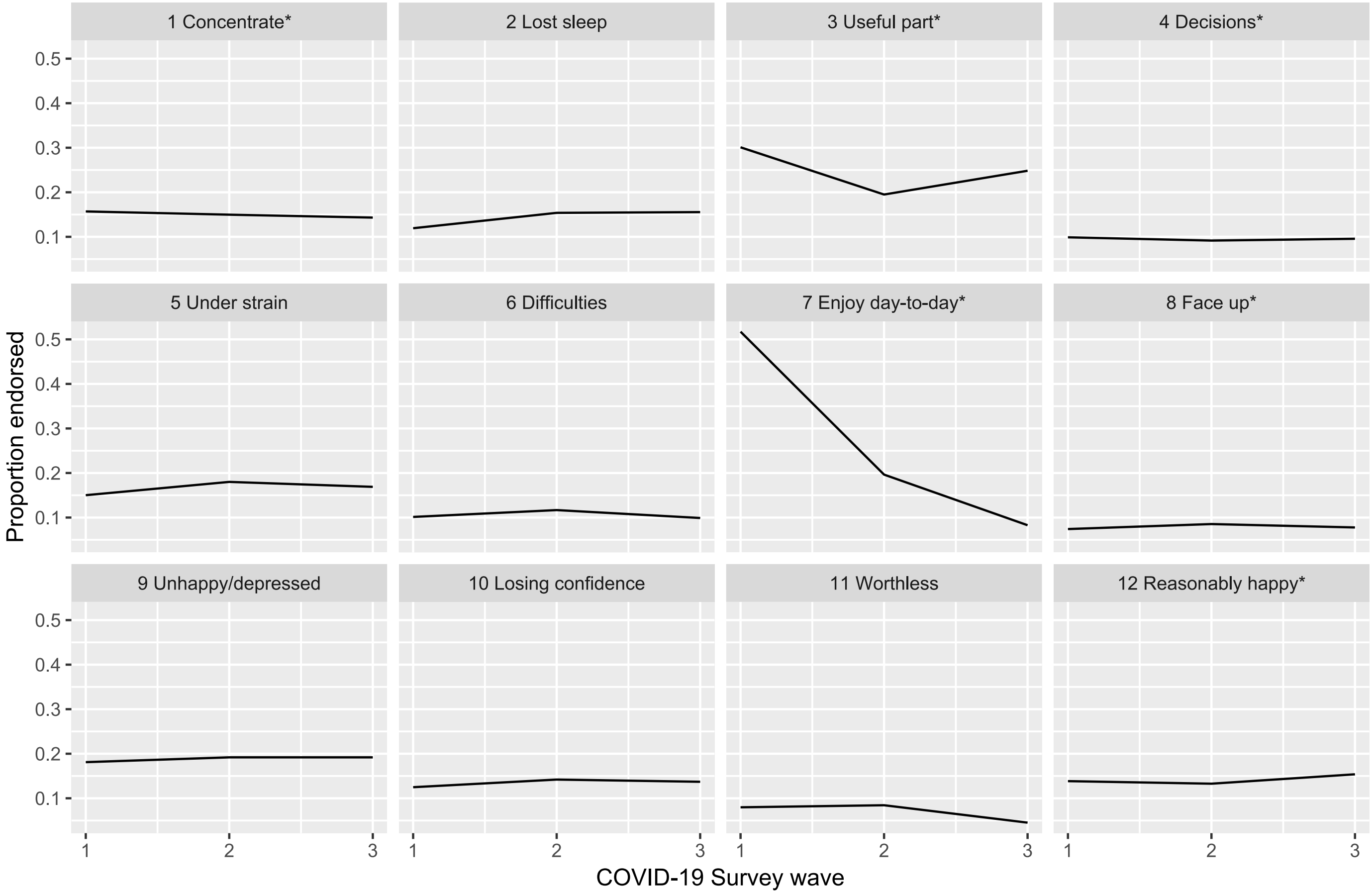

**eTable 3.1. Weighted prevalence estimates (and 95% CIs) of psychological distress 'caseness' across COVID-19 Survey waves by operationalisation**

| | Operationalisation A, proportion (95% CI) | Operationalisation B, proportion (95% CI) | Operationalisation C, proportion (95% CI) |
|---|---|---|---|
| Survey wave 1 | 0.41 [0.39, 0.44] | 0.32 [0.30, 0.34] | 0.49 [0.47, 0.52] |
| Survey wave 2 | 0.39 [0.37, 0.41] | 0.36 [0.34, 0.38] | 0.52 [0.50, 0.54] |
| Survey wave 3 | 0.29 [0.27, 0.32] | 0.28 [0.26, 0.30] | 0.42 [0.40, 0.44] |

*Note*. Confidence intervals (CIs) are constructed based on the Agresti-Coull method. Operationalisation A: keeping item 7 and threshold at 2+; Operationalisation B: dropping item 7 and keeping threshold at 2+; Operationalisation C: dropping item 7 and lowering threshold to 1+.

### eAppendix 4. Measurement invariance testing

As comparisons of the psychological distress' levels were going to be made across time points, cohorts (in the case of the models estimated using the same indicators), and sexes (in the case of the stratified models), we explored the degree of measurement invariance across these conditions where the same set of indicators were going to be used.[10]

We extended the existing evidence on the invariant properties of the Malaise Inventory over time, across cohorts, and sexes,[11] to include the most recent data collection points. We implemented the same procedure with the GAD-2 and PHQ-2 items to assess if they had equivalent measurement properties across the three cohorts, time-points, and sexes, as they were used as 'anchor items' in the factor score approach including the three cohorts.

More specifically, the level of invariance needed to make meaningful comparisons across conditions was different for the models using different operationalisations.

The most restrictive level of invariance was needed for the models estimated as a sensitivity check using the number of psychological distress symptoms according to the Malaise Inventory, used in the NCDS and BCS70 cohorts, as comparisons were going to be made at the observed number of symptoms (count) level, where all symptoms had the same 'weight'. Thus, not only the item loadings and thresholds had to be equivalent across conditions (scalar/strong invariance), but also the loadings should be the same across items (Rasch-type model), as each of the items were counted as a unit. Evidence of this level of invariance across time points, cohorts, and sexes had been previously obtained up to the most recent pre-pandemic assessment,[9,11] and we extended that analysis to include the three COVID-19 Survey waves.

In the factor score approach with the three cohorts, a number of indicators common across the three cohorts ('anchor' items, the GAD-2 and PHQ-2 items) were going to be used alongside the cohort- and wave-specific indicators to measure a latent psychological distress factor. Unlike the previous case, in this approach the scalar/strong invariance level would suffice, as comparisons were going to be made across factor scores, which had been obtained based on the different 'weights' (loadings) of each of the items. In this case, however, the measurement invariance testing would be limited to the 'anchor' items as a way of exploring to what extent this subset of common items was comparable across conditions (time points, cohorts, and sexes).

A structural equation modelling (SEM) framework was used, where the psychological distress latent factors were defined by their corresponding set of indicators (the nine items of the Malaise Inventory, or the GAD-2 and PHQ-2 items). Due to the nature of the indicators (categorical ordered), models were estimated using a Weighted Least Squares Mean and Variance adjusted (WLSMV) estimator and under a Delta parameterisation.[12]

Models with increasing levels of constraints were estimated.[10] First, models where the same factor structure was proposed across conditions without further constraints (configural invariance model) were estimated. The fit of these configural models to the data was assessed by means of the goodness-of-fit indices, with Root Mean Square Error of Approximation (RMSEA) values close to 0.060, and Comparative Fit Index (CFI) and Tucker-Lewis Index values close to 0.950 being generally considered as indicative of good fit.[13] Then, models where both the items' loadings and thresholds were constrained to be equal across the different conditions (scalar/strong invariance model) were estimated. The goodness-of-fit indices of these models were compared with those from the configural models and deemed to be invariant (i.e., to not fit the data substantially worse) if the loss in fit was smaller than 0.010 for the CFI and 0.015 for the RMSEA.[14,15] Finally, and only for the models with the Malaise Inventory, additional models were estimated where all items were constrained to have the same factor loading (Rasch-type model).

Based on the abovementioned criteria, scalar invariance was found to hold for the Malaise inventory in all cases tested **eTable 4.1**). The further constrained Rasch-type model was found to fit well the data.

In the case of the GAD-2 and PHQ-2 items, RMSEA values were found to be beyond the usual threshold for acceptable fit, whereas the CFI and TLI indices suggested good fit across the different models (**eTable 4.2**). Due to the sensitivity of the RMSEA fit index to models with small degrees of freedom [16] (in this case, 2 degrees of freedom per group in the configural models), and considering the good performance in the alternative indices and the improvement in comparative fit with increasing constraints, we considered scalar invariance to hold.

**eTable 4.1. Malaise inventory measurement invariance testing results**

| | Model | Chi-square (df) | RMSEA (90% CI) | CFI | TLI | ΔRMSEA | ΔCFI |
|---|---|---|---|---|---|---|---|
| NCDS: age (7: 23, 33, 42, 50, 61.7, 62, 62.5, 63) * birth sex (2) | Configural | 1807 (432) | 0.034 (0.032, 0.035) | 0.987 | 0.983 | | |
| | Scalar | 2837 (537) | 0.039 (0.038, 0.040) | 0.978 | 0.977 | -0.005 | -0.009 |
| | **Rasch** | **3170 (545)** | **0.041 (0.040, 0.043)** | **0.975** | **0.974** | **-0.002** | **-0.003** |
| BCS70: age (8: 26, 29, 34, 42, 46, 50, 50.5, 51) * birth sex (2) | Configural | 2333 (432) | 0.041 (0.039, 0.043) | 0.985 | 0.980 | | |
| | Scalar | 2962 (537) | 0.041 (0.040, 0.043) | 0.981 | 0.979 | <0.001 | -0.004 |
| | **Rasch** | **3424 (545)** | **0.045 (0.043, 0.046)** | **0.977** | **0.976** | **-0.004** | **-0.004** |
| NCDS/BCS70: age (4: 23/26, 33/34, 42, 50) * birth sex (2) * cohort (2) | Configural | 2028 (432) | 0.036 (0.034, 0.037) | 0.985 | 0.980 | | |
| | Scalar | 2964 (537) | 0.039 (0.038, 0.041) | 0.977 | 0.975 | -0.003 | -0.008 |
| | **Rasch** | **3205 (545)** | **0.041 (0.040, 0.042)** | **0.975** | **0.973** | **-0.002** | **-0.002** |

*Note*. BCS70: 1970 British Cohort Study; CFI: Comparative Fit Index; df: degrees of freedom; NCDS: 1958 National Child Development Study; NSHD: 1946 National Survey of Health and Development; RMSEA: Root Mean Square Error of Approximation; TLI: Tucker-Lewis Index; ΔCFI: difference in CFI; ΔRMSEA: difference in RMSEA. Selected models are highlighted in bold.

**eTable 4.2. GAD-2 and PHQ-2 measurement invariance testing results**

| | Model | Chi-square (df) | RMSEA (90% CI) | CFI | TLI | ΔRMSEA | ΔCFI |
|---|---|---|---|---|---|---|---|
| NSHD: CWs (3) * birth sex (2) | Configural | 384 (12) | 0.183 (0.167, 0.199) | 0.991 | 0.974 | | |
| | **Scalar** | **553 (62)** | **0.092 (0.085, 0.100)** | **0.989** | **0.993** | **0.091** | **-0.002** |
| NCDS: CWs (3) * birth sex (2) | Configural | 1402 (12) | 0.200 (0.191, 0.209) | 0.987 | 0.960 | | |
| | **Scalar** | **1881 (62)** | **0.100 (0.097, 0.104)** | **0.983** | **0.990** | **0.100** | **-0.004** |
| BCS70: CWs (3) * birth sex (2) | Configural | 1494 (12) | 0.227 (0.218, 0.237) | 0.985 | 0.955 | | |
| | **Scalar** | **2026 (62)** | **0.115 (0.111, 0.120)** | **0.980** | **0.989** | **0.112** | **-0.005** |
| NSHD+NCDS+BCS70: CWs (3) * birth sex (2) * cohort (3) | Configural | 3289 (36) | 0.209 (0.203, 0.215) | 0.987 | 0.961 | | |
| | **Scalar** | **4759 (206)** | **0.103 (0.101, 0.106)** | **0.982** | **0.990** | **0.106** | **-0.005** |

*Note*. BCS70: 1970 British Cohort Study; CWs: COVID-19 Survey waves. CFI: Comparative Fit Index; df: degrees of freedom; NCDS: 1958 National Child Development Study; NSHD: 1946 National Survey of Health and Development; RMSEA: Root Mean Square Error of Approximation; TLI: Tucker-Lewis Index; ΔCFI: difference in CFI; ΔRMSEA: difference in RMSEA. Selected models are highlighted in bold.

**eAppendix 5. SEM diagram for the measurement model used in the factor score approach with the three cohorts (NSHD, NCDS, and BCS70)**

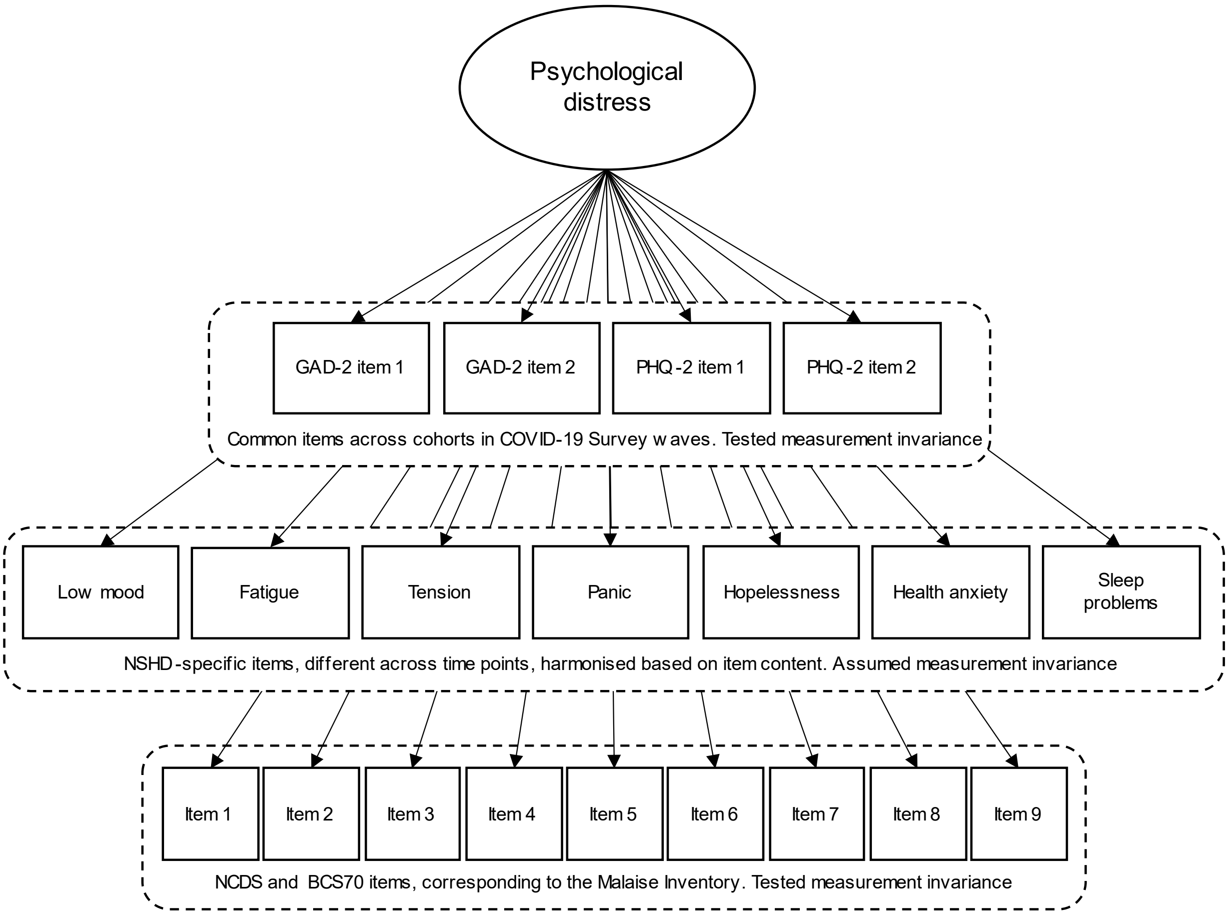

Note. BCS70: 1970 British Cohort Study; GAD-2: 2-item Generalised Anxiety Disorder questionnaire; NCDS: 1958 National Child Development Study; NSHD: 1946 National Survey of Health and Development; PHQ-2: 2-item Patient Health Questionnaire.

**eAppendix 6. Details on the analytical approach used to model distress trajectories and to obtain projections of the distress levels "had the pandemic not occurred"**

We used a multilevel growth curve modelling approach to analyse the trajectories of psychological distress under the different outcome operationalisations, using linear models for the factor scores operationalisations (continuous), Poisson models for the number of symptoms operationalisation (discrete), and logistic models for the 'caseness' operationalisation (binary). To model the non-linear trajectories observed in the descriptive data, we used a piecewise approach with two main segments.

The first segment covered the period from the first time-point to the last pre-pandemic assessment and corresponded to the functional form reported in the previous study for this period,[8] which was quadratic (inverted U-pattern) for NSHD and cubic (U-pattern followed by a decrease or stabilisation) for BCS70. An additional polynomial term (quartic) was included in NCDS to model a slight increase in the trajectory towards the last pre-pandemic assessment. The second segment covered the period from the last pre-pandemic assessment to the study period in February/March 2021 and was defined by a polynomial curve up to the cubic term to capture the observed multifaceted change.

Unadjusted models were estimated separately for each cohort. The models were also estimated including an interaction term between each growth parameter and birth sex, to account for inequalities in these trajectories within cohorts in line with the abovementioned evidence. The random part of all these models included the variation in the initial levels (random intercepts) but not in the change over time (random slopes) as the inclusion of this additional random effect led to convergence issues.

To answer the counterfactual question of what the distress levels would have been had the COVID-19 pandemic not occurred, models were estimated using data only up to the most recent pre-pandemic assessment.

The same models used when including the data from the COVID-19 Survey waves were not rendered useful for obtaining projections, as the polynomial terms produced unlikely predictions. Therefore, a piecewise approach with two segments was used, locating the knot at the middle point of the pre-pandemic trajectory in order to maximise the data available to estimate each of the two segments. At least three time points per segment were necessary to enable the estimation of non-linear trajectories in each of the segments; this is, a minimum total number of five observations, with the first to the third belonging to the first segment, and the third to the fifth belonging to the second segment.

The models were estimated separately for each cohort using the main psychological distress operationalisation (cross-cohort factor score). The segments comprised years 1982, 1989, and 1999 (first segment), and 1999, 2009, and 2015 (second segment) in NSHD; years 1981, 1991, and 2000 (first segment), and 2000, 2008, and 2020 (second segment) in NCDS; and years 1996, 1999, and 2004 (first segment), and 2004, 2012, and 2016 (second segment) in BCS70.

These models were used to obtain 95% confidence intervals of the mean psychological distress factor score in 2020 and 2021. These confidence intervals were plotted against those obtained from the models estimated using the complete data (this is, also including data from the COVID-19 Survey waves).

**eAppendix 7. Sample flow diagram**

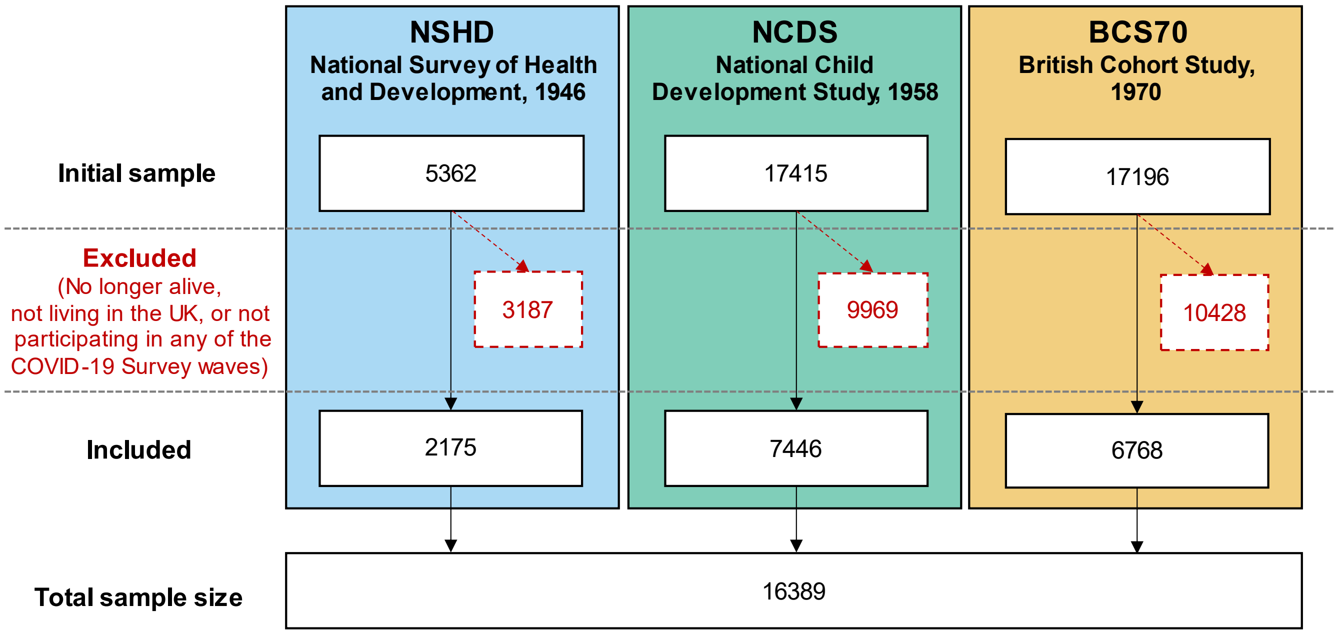

### eAppendix 8. Results from multilevel growth curve models with cross-cohort factor scores as outcome (linear models)

**eTable 8.1. Model coefficients from the multilevel growth curve models with cross-cohort factor scores as outcome (linear models)**

| | NSHD | | NCDS | | BCS70 | |
|---|---|---|---|---|---|---|
| **Models without interaction by birth sex** | **Coefficient (95% CI)** | ***p*** | **Coefficient (95% CI)** | ***p*** | **Coefficient (95% CI)** | ***p*** |
| Spline 1, linear term | 0.02 (0.02, 0.02) | <0.001 | -0.10 (-0.11, -0.09) | <0.001 | -0.06 (-0.07, -0.05) | <0.001 |
| Spline 1, quadratic term | -0.001 (-0.001, 0.000) | <0.001 | 0.013 (0.012, 0.014) | <0.001 | 0.007 (0.007, 0.008) | <0.001 |
| Spline 1, cubic term | | | -0.0005 (-0.0006, -0.0005) | <0.001 | -0.0002 (-0.0003, -0.0002) | <0.001 |
| Spline 1, quartic term | | | 0.00001 (0.00001, 0.00001) | <0.001 | | |
| Spline 2, linear term | -1.17 (-2.58, 0.24) | 0.105 | 0.04 (-0.15, 0.23) | 0.664 | -0.47 (-0.92, -0.02) | 0.041 |
| Spline 2, quadratic term | 0.43 (-0.09, 0.95) | 0.104 | 0.11 (-0.22, 0.45) | 0.511 | 0.22 (0.02, 0.42) | 0.031 |
| Spline 2, cubic term | -0.04 (-0.09, 0.01) | 0.111 | -0.08 (-0.24, 0.08) | 0.343 | -0.02 (-0.05, 0.00) | 0.033 |
| Intercept | -0.14 (-0.17, -0.11) | <0.001 | -0.17 (-0.19, -0.16) | <0.001 | 0.12 (0.10, 0.14) | <0.001 |
| Intercept variance | 0.20 (0.18, 0.22) | | 0.34 (0.33, 0.35) | | 0.40 (0.38, 0.41) | |
| **Models with interaction by birth sex** | **Coefficient (95% CI)** | ***p*** | **Coefficient (95% CI)** | ***p*** | **Coefficient (95% CI)** | ***p*** |
| Spline 1, linear term | 0.02 (0.01, 0.03) | <0.001 | -0.08 (-0.09, -0.07) | <0.001 | -0.04 (-0.05, -0.03) | <0.001 |
| Spline 1, quadratic term | -0.001 (-0.001, 0.000) | <0.001 | 0.012 (0.010, 0.013) | <0.001 | 0.006 (0.005, 0.007) | <0.001 |
| Spline 1, cubic term | | | -0.0005 (-0.0005, -0.0004) | <0.001 | -0.0002 (-0.0002, -0.0002) | <0.001 |
| Spline 1, quartic term | | | 0.00001 (0.00001, 0.00001) | <0.001 | | |
| Spline 2, linear term | -2.03 (-4.15, 0.10) | 0.062 | -0.32 (-0.58, -0.05) | 0.020 | -0.38 (-1.09, 0.33) | 0.291 |
| Spline 2, quadratic term | 0.73 (-0.06, 1.51) | 0.070 | 0.55 (0.08, 1.03) | 0.021 | 0.17 (-0.14, 0.49) | 0.279 |
| Spline 2, cubic term | -0.06 (-0.14, 0.01) | 0.078 | -0.24 (-0.47, -0.02) | 0.033 | -0.02 (-0.05, 0.02) | 0.289 |
| Intercept * women | 0.14 (0.08, 0.20) | <0.001 | 0.39 (0.36, 0.42) | <0.001 | 0.35 (0.31, 0.39) | <0.001 |
| Spline 1, linear term * women | 0.00 (-0.01, 0.01) | 0.900 | -0.03 (-0.04, -0.01) | <0.001 | -0.03 (-0.05, -0.02) | <0.001 |
| Spline 1, quadratic term * women | 0.000 (0.000, 0.000) | 0.986 | 0.002 (0.000, 0.004) | 0.052 | 0.002 (0.001, 0.004) | 0.008 |
| Spline 1, cubic term * women | | | -0.0001 (-0.0001, 0.0000) | 0.209 | -0.0001 (-0.0001, 0.0000) | 0.068 |
| Spline 1, quartic term * women | | | 0.00000 (0.00000, 0.00000) | 0.397 | | |
| Spline 2, linear term * women | 1.65 (-1.16, 4.46) | 0.249 | 0.70 (0.32, 1.08) | <0.001 | -0.15 (-1.06, 0.76) | 0.748 |
| Spline 2, quadratic term * women | -0.57 (-1.61, 0.46) | 0.279 | -0.87 (-1.55, -0.20) | 0.011 | 0.08 (-0.32, 0.48) | 0.701 |
| Spline 2, cubic term * women | 0.05 (-0.04, 0.14) | 0.301 | 0.33 (0.01, 0.65) | 0.043 | -0.01 (-0.05, 0.04) | 0.690 |
| Intercept | -0.21 (-0.25, -0.17) | <0.001 | -0.38 (-0.40, -0.36) | <0.001 | -0.08 (-0.11, -0.05) | <0.001 |

| | | | |
|---|---|---|---|
| Intercept variance | 0.19 (0.17, 0.21) | 0.31 (0.30, 0.32) | 0.38 (0.37, 0.39) |

*Note.* Unadjusted results. BCS70: 1970 British Cohort Study; NCDS: 1958 National Child Development Study; NSHD: 1946 National Survey of Health and Development.

**eTable 8.2. Marginal mean levels predicted from the multilevel growth curve models with cross-cohort factor scores as outcome (linear models)**

| | | | All | Men | Women |
|---|---|---|---|---|---|
| | **Age** | **Year** | **Marginal mean (95% CI)** | **Marginal mean (95% CI)** | **Marginal mean (95% CI)** |
| **NSHD** | 36 | 1982 | -0.14 (-0.17, -0.11) | -0.21 (-0.25, -0.17) | -0.07 (-0.12, -0.03) |
| | 43 | 1989 | -0.03 (-0.05, 0.00) | -0.10 (-0.14, -0.07) | 0.04 (0.00, 0.07) |
| | 53 | 1999 | 0.02 (-0.01, 0.05) | -0.06 (-0.10, -0.01) | 0.09 (0.05, 0.13) |
| | 63 | 2009 | -0.06 (-0.08, -0.03) | -0.14 (-0.17, -0.10) | 0.02 (-0.02, 0.06) |
| | 69 | 2015 | -0.16 (-0.19, -0.13) | -0.24 (-0.29, -0.20) | -0.09 (-0.13, -0.04) |
| | 74 | May 2020 | -0.08 (-0.13, -0.04) | -0.28 (-0.34, -0.22) | 0.10 (0.04, 0.16) |
| | 74.5 | Sept/Oct 2020 | -0.01 (-0.07, 0.05) | -0.15 (-0.23, -0.07) | 0.12 (0.05, 0.20) |
| | 75 | Feb/Mar 2021 | -0.04 (-0.10, 0.02) | -0.19 (-0.28, -0.10) | 0.10 (0.02, 0.19) |
| | **Age** | **Year** | **Marginal mean (95% CI)** | **Marginal mean (95% CI)** | **Marginal mean (95% CI)** |
| **NCDS** | 23 | 1981 | -0.17 (-0.19, -0.16) | -0.38 (-0.40, -0.36) | 0.01 (-0.01, 0.03) |
| | 33 | 1991 | -0.31 (-0.32, -0.29) | -0.44 (-0.46, -0.42) | -0.18 (-0.21, -0.16) |
| | 42 | 2000 | -0.04 (-0.06, -0.02) | -0.18 (-0.21, -0.16) | 0.09 (0.07, 0.12) |
| | 50 | 2008 | -0.08 (-0.10, -0.06) | -0.23 (-0.26, -0.20) | 0.05 (0.02, 0.08) |
| | 61.7 | Jan/Mar 2020 | -0.06 (-0.09, -0.03) | -0.18 (-0.23, -0.13) | 0.05 (0.00, 0.09) |
| | 62 | May 2020 | -0.04 (-0.06, -0.01) | -0.23 (-0.27, -0.20) | 0.14 (0.11, 0.18) |
| | 62.5 | Sept/Oct 2020 | 0.01 (-0.02, 0.03) | -0.20 (-0.23, -0.16) | 0.20 (0.16, 0.23) |
| | 63 | Feb/Mar 2021 | 0.01 (-0.01, 0.04) | -0.19 (-0.22, -0.16) | 0.20 (0.17, 0.23) |
| | **Age** | **Year** | **Marginal mean (95% CI)** | **Marginal mean (95% CI)** | **Marginal mean (95% CI)** |
| **BCS70** | 26 | 1996 | 0.12 (0.10, 0.14) | -0.08 (-0.11, -0.05) | 0.27 (0.25, 0.30) |
| | 29 | 1999 | 0.01 (-0.01, 0.02) | -0.15 (-0.17, -0.12) | 0.12 (0.10, 0.15) |
| | 34 | 2004 | 0.01 (-0.01, 0.03) | -0.11 (-0.14, -0.08) | 0.10 (0.08, 0.13) |
| | 42 | 2012 | 0.14 (0.12, 0.16) | 0.02 (-0.01, 0.05) | 0.23 (0.20, 0.26) |
| | 46 | 2016 | 0.07 (0.04, 0.09) | -0.05 (-0.09, -0.02) | 0.16 (0.13, 0.19) |
| | 50 | May 2020 | 0.17 (0.14, 0.19) | 0.00 (-0.04, 0.04) | 0.30 (0.27, 0.34) |
| | 50.5 | Sept/Oct 2020 | 0.21 (0.19, 0.24) | 0.04 (0.00, 0.08) | 0.36 (0.32, 0.39) |
| | 51 | Feb/Mar 2021 | 0.21 (0.18, 0.23) | 0.04 (-0.01, 0.08) | 0.35 (0.31, 0.38) |

*Note.* Unadjusted results. BCS70: 1970 British Cohort Study; NCDS: 1958 National Child Development Study; NSHD: 1946 National Survey of Health and Development.

**eFigure 8.1. Marginal mean cross-cohort psychological distress factor scores over time (year and age) by birth sex**

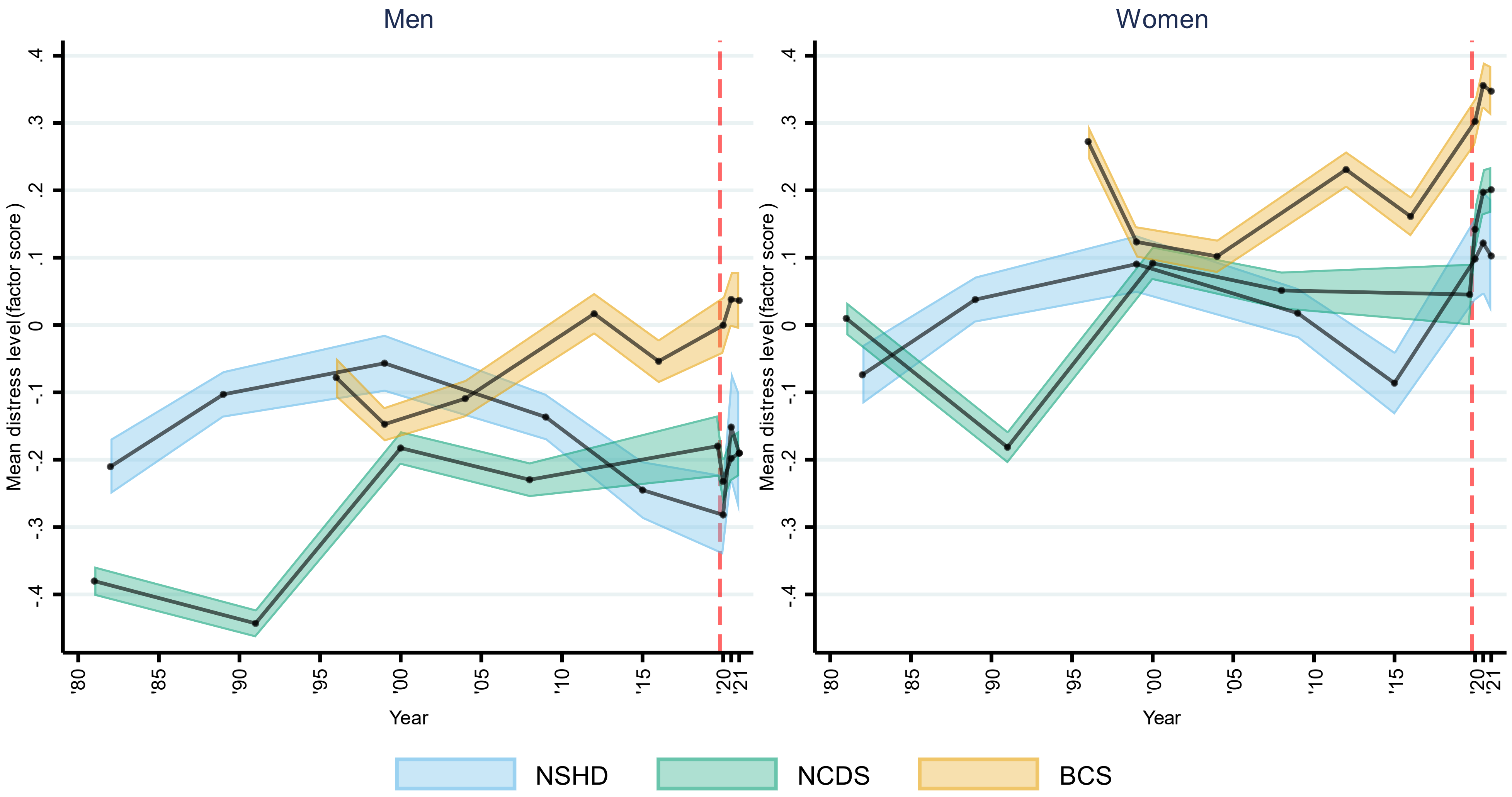

Note: Unadjusted results . 95% confidence intervals are indicated in lighter shaded areasBCS70: 1970 British Cohort Study, NCDS: 1958 National Child and Development Study; NSHD: 1946 National Survey of Health and Development .
The dashed line represents the first nationwide lockdown enforced in March 2020.

### eAppendix 9. Results from multilevel growth curve models with number of symptoms as outcome (Poisson models)

**eTable 9.1. Model coefficients from the multilevel growth curve models with number of symptoms as outcome (Poisson models)**

| | NSHD | | NCDS | | BCS70 | |
|---|---|---|---|---|---|---|
| **Models without interaction by birth sex** | **Coefficient (95% CI)** | ***p*** | **Coefficient (95% CI)** | ***p*** | **Coefficient (95% CI)** | ***p*** |
| Spline 1, linear term | 0.05 (0.03, 0.06) | <0.001 | -0.17 (-0.18, -0.15) | <0.001 | -0.06 (-0.07, -0.05) | <0.001 |
| Spline 1, quadratic term | -0.002 (-0.002, -0.001) | <0.001 | 0.022 (0.020, 0.024) | <0.001 | 0.008 (0.007, 0.009) | <0.001 |
| Spline 1, cubic term | | | -0.0008 (-0.0009, -0.0008) | <0.001 | -0.0003 (-0.0003, -0.0002) | <0.001 |
| Spline 1, quartic term | | | 0.00001 (0.00001, 0.00001) | <0.001 | | |
| Spline 2, linear term | -1.31 (-7.14, 4.52) | 0.659 | -0.35 (-0.65, -0.04) | 0.025 | -1.86 (-2.38, -1.34) | <0.001 |
| Spline 2, quadratic term | 0.52 (-1.64, 2.68) | 0.635 | 1.08 (0.56, 1.59) | <0.001 | 0.83 (0.60, 1.06) | <0.001 |
| Spline 2, cubic term | -0.05 (-0.25, 0.15) | 0.639 | -0.59 (-0.82, -0.35) | <0.001 | -0.09 (-0.12, -0.07) | <0.001 |
| Intercept | -1.70 (-1.82, -1.59) | <0.001 | -0.44 (-0.48, -0.40) | <0.001 | 0.13 (0.10, 0.16) | <0.001 |
| Intercept variance | 1.60 (1.44, 1.77) | | 1.24 (1.18, 1.30) | | 0.96 (0.91, 1.00) | |
| **Models with interaction by birth sex** | **Coefficient (95% CI)** | ***p*** | **Coefficient (95% CI)** | ***p*** | **Coefficient (95% CI)** | ***p*** |
| Spline 1, linear term | 0.06 (0.04, 0.08) | <0.001 | -0.19 (-0.22, -0.16) | <0.001 | -0.05 (-0.07, -0.03) | <0.001 |
| Spline 1, quadratic term | -0.002 (-0.003, -0.001) | <0.001 | 0.027 (0.023, 0.030) | <0.001 | 0.008 (0.005, 0.010) | <0.001 |
| Spline 1, cubic term | | | -0.0011 (-0.0012, -0.0009) | <0.001 | -0.0002 (-0.0003, -0.0002) | <0.001 |
| Spline 1, quartic term | | | 0.00001 (0.00001, 0.00002) | <0.001 | | <0.001 |
| Spline 2, linear term | -8.51 (-14.66, -2.36) | 0.007 | -0.56 (-1.07, -0.06) | 0.029 | -1.67 (-2.64, -0.69) | 0.001 |
| Spline 2, quadratic term | 3.16 (0.89, 5.42) | 0.006 | 1.27 (0.39, 2.15) | 0.005 | 0.73 (0.30, 1.17) | 0.001 |
| Spline 2, cubic term | -0.29 (-0.50, -0.08) | 0.007 | -0.64 (-1.05, -0.23) | 0.002 | -0.08 (-0.13, -0.03) | 0.001 |
| Intercept * women | 0.53 (0.32, 0.74) | <0.001 | 0.90 (0.82, 0.97) | <0.001 | 0.57 (0.51, 0.64) | <0.001 |
| Spline 1, linear term * women | -0.02 (-0.05, 0.01) | 0.176 | 0.04 (0.00, 0.07) | 0.036 | -0.02 (-0.05, 0.00) | 0.036 |
| Spline 1, quadratic term * women | 0.001 (0.000, 0.002) | 0.077 | -0.008 (-0.012, -0.004) | <0.001 | 0.001 (-0.002, 0.003) | 0.557 |
| Spline 1, cubic term * women | | | 0.0004 (0.0002, 0.0005) | <0.001 | 0.0000 (-0.0001, 0.0001) | 0.982 |
| Spline 1, quartic term * women | | | 0.00000 (-0.00001, 0.00000) | <0.001 | | <0.001 |
| Spline 2, linear term * women | 11.06 (1.12, 21.00) | 0.029 | 0.37 (-0.26, 1.00) | 0.253 | -0.26 (-1.40, 0.87) | 0.651 |
| Spline 2, quadratic term * women | -4.05 (-7.72, -0.37) | 0.031 | -0.34 (-1.42, 0.75) | 0.546 | 0.13 (-0.37, 0.64) | 0.601 |
| Spline 2, cubic term * women | 0.37 (0.03, 0.71) | 0.032 | 0.10 (-0.41, 0.61) | 0.700 | -0.02 (-0.07, 0.04) | 0.583 |
| Intercept | -1.98 (-2.16, -1.80) | <0.001 | -0.95 (-1.02, -0.88) | <0.001 | -0.20 (-0.26, -0.15) | <0.001 |
| Intercept variance | 1.51 (1.36, 1.67) | | 1.13 (1.08, 1.18) | | 0.90 (0.86, 0.95) | |

*Note.* Unadjusted results. BCS70: 1970 British Cohort Study; NCDS: 1958 National Child Development Study; NSHD: 1946 National Survey of Health and Development.

**eTable 9.2. Marginal mean levels predicted from the multilevel growth curve models with number of symptoms as outcome (Poisson models)**

| | Age | Year | Marginal mean (95% CI) |
|---|---|---|---|
| NSHD | 36 | 1982 | 0.40 (0.37, 0.44) |
| | 43 | 1989 | 0.52 (0.48, 0.55) |
| | 53 | 1999 | 0.55 (0.51, 0.60) |
| | 63 | 2009 | 0.42 (0.39, 0.46) |
| | 69 | 2015 | 0.31 (0.27, 0.34) |
| | 74 | May 2020 | 0.54 (0.48, 0.60) |
| | 74.5 | Sept/Oct 2020 | 0.60 (0.49, 0.72) |
| | 75 | Feb/Mar 2021 | 0.60 (0.47, 0.73) |
| | **Age** | **Year** | **Marginal mean (95% CI)** |
| NCDS | 23 | 1981 | 1.19 (1.15, 1.23) |
| | 33 | 1991 | 0.96 (0.92, 1.00) |
| | 42 | 2000 | 1.55 (1.51, 1.60) |
| | 50 | 2008 | 1.51 (1.46, 1.56) |
| | 61.7 | Jan/Mar 2020 | 1.51 (1.42, 1.59) |
| | 62 | May 2020 | 1.48 (1.42, 1.54) |
| | 62.5 | Sept/Oct 2020 | 1.70 (1.64, 1.76) |
| | 63 | Feb/Mar 2021 | 1.61 (1.55, 1.67) |
| | **Age** | **Year** | **Marginal mean (95% CI)** |
| BCS70 | 26 | 1996 | 1.84 (1.79, 1.89) |
| | 29 | 1999 | 1.63 (1.58, 1.67) |
| | 34 | 2004 | 1.65 (1.60, 1.69) |
| | 42 | 2012 | 1.95 (1.89, 2.00) |
| | 46 | 2016 | 1.85 (1.79, 1.90) |
| | 50 | May 2020 | 1.88 (1.81, 1.94) |
| | 50.5 | Sept/Oct 2020 | 2.14 (2.08, 2.21) |
| | 51 | Feb/Mar 2021 | 2.00 (1.94, 2.07) |

*Note.* Unadjusted results. BCS70: 1970 British Cohort Study; NCDS: 1958 National Child Development Study; NSHD: 1946 National Survey of Health and Development.

**eFigure 9.1. Marginal mean number of psychological distress symptoms over time (year and age)**

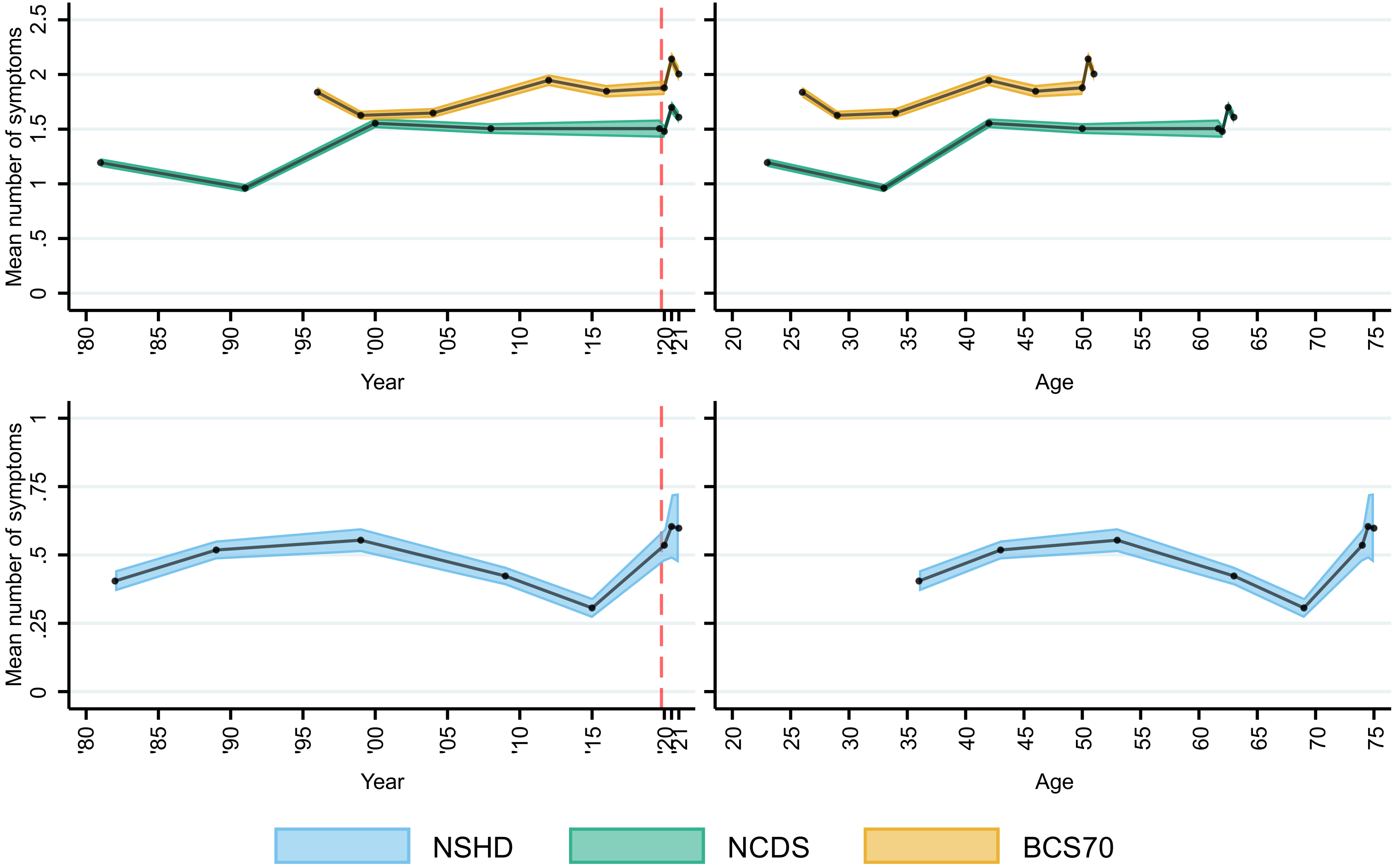

Note: Unadjusted results. 95% confidence intervals are indicated in lighter shaded areas . BCS70: 1970 British Cohort Study; NCDS: 1958 National Child and Development Study; NSHD: 1946 National Survey of Health and Development . Y axes are different for NCDS/BCS70 and NSHD as they reflect different total possible numbers of symptoms .
The dashed line represents the first nationwide lockdown enforced in March 2020.

### eAppendix 10. Results from multilevel growth curve models with caseness as outcome (logistic models)

**eTable 10.1. Model coefficients from the multilevel growth curve models with caseness as outcome (logistic models)**

| | NSHD | | NCDS | | BCS70 | |
|---|---|---|---|---|---|---|
| **Models without interaction by birth sex** | **Coefficient (95% CI)** | ***p*** | **Coefficient (95% CI)** | ***p*** | **Coefficient (95% CI)** | ***p*** |
| Spline 1, linear term | 0.18 (0.14, 0.21) | <0.001 | -0.30 (-0.38, -0.23) | <0.001 | -0.14 (-0.19, -0.08) | <0.001 |
| Spline 1, quadratic term | -0.004 (-0.005, -0.004) | <0.001 | 0.039 (0.029, 0.049) | <0.001 | 0.017 (0.011, 0.024) | <0.001 |
| Spline 1, cubic term | 0.00 (0.00, 0.00) | <0.001 | -0.0014 (-0.0018, -0.0010) | <0.001 | -0.0005 (-0.0007, -0.0003) | <0.001 |
| Spline 1, quartic term | 0.00 (0.00, 0.00) | <0.001 | 0.00002 (0.00001, 0.00002) | <0.001 | 0.00 (0.00, 0.00) | <0.001 |
| Spline 2, linear term | -2.60 (-13.45, 8.24) | 0.638 | -1.62 (-3.06, -0.18) | 0.028 | -5.68 (-8.30, -3.07) | <0.001 |
| Spline 2, quadratic term | 1.21 (-2.80, 5.22) | 0.555 | 4.08 (1.61, 6.54) | 0.001 | 2.50 (1.34, 3.66) | <0.001 |
| Spline 2, cubic term | -0.12 (-0.49, 0.25) | 0.521 | -2.06 (-3.20, -0.92) | <0.001 | -0.27 (-0.40, -0.14) | <0.001 |
| Intercept | -4.52 (-4.86, -4.19) | <0.001 | -4.69 (-4.89, -4.49) | <0.001 | -3.27 (-3.41, -3.12) | <0.001 |
| Intercept variance | 5.05 (4.44, 5.75) | | 7.66 (6.94, 8.45) | | 6.87 (6.32, 7.47) | |
| **Models with interaction by birth sex** | **Coefficient (95% CI)** | ***p*** | **Coefficient (95% CI)** | ***p*** | **Coefficient (95% CI)** | ***p*** |
| Spline 1, linear term | 0.20 (0.15, 0.26) | <0.001 | -0.32 (-0.47, -0.18) | <0.001 | -0.11 (-0.21, -0.02) | 0.017 |
| Spline 1, quadratic term | -0.005 (-0.007, -0.004) | <0.001 | 0.049 (0.031, 0.067) | <0.001 | 0.018 (0.008, 0.029) | 0.001 |
| Spline 1, cubic term | 0.00 (0.00, 0.00) | <0.001 | -0.0019 (-0.0026, -0.0012) | <0.001 | -0.0005 (-0.0009, -0.0002) | 0.002 |
| Spline 1, quartic term | 0.00 (0.00, 0.00) | <0.001 | 0.00002 (0.00001, 0.00003) | <0.001 | 0.00 (0.00, 0.00) | <0.001 |
| Spline 2, linear term | -17.57 (-30.64, -4.51) | 0.008 | -2.91 (-5.26, -0.56) | 0.015 | -5.26 (-9.99, -0.54) | 0.029 |
| Spline 2, quadratic term | 6.70 (1.88, 11.53) | 0.006 | 5.22 (1.15, 9.30) | 0.012 | 2.27 (0.18, 4.36) | 0.034 |
| Spline 2, cubic term | -0.62 (-1.07, -0.18) | 0.006 | -2.37 (-4.27, -0.46) | 0.015 | -0.24 (-0.47, -0.01) | 0.038 |
| Intercept * women | 1.29 (0.68, 1.91) | <0.001 | 2.02 (1.68, 2.36) | <0.001 | 1.23 (0.97, 1.50) | <0.001 |
| Spline 1, linear term * women | -0.04 (-0.12, 0.03) | 0.237 | 0.04 (-0.13, 0.21) | 0.621 | -0.03 (-0.14, 0.08) | 0.599 |
| Spline 1, quadratic term * women | 0.001 (-0.001, 0.003) | 0.162 | -0.016 (-0.037, 0.005) | 0.141 | -0.002 (-0.015, 0.011) | 0.769 |
| Spline 1, cubic term * women | 0.00 (0.00, 0.00) | <0.001 | 0.0008 (-0.0001, 0.0017) | 0.071 | 0.0001 (-0.0003, 0.0005) | 0.657 |
| Spline 1, quartic term * women | 0.00 (0.00, 0.00) | <0.001 | -0.00001 (-0.00002, 0.00000) | 0.047 | 0.00 (0.00, 0.00) | <0.001 |
| Spline 2, linear term * women | 26.80 (7.61, 45.98) | 0.006 | 2.15 (-0.82, 5.12) | 0.156 | -0.65 (-6.27, 4.98) | 0.822 |
| Spline 2, quadratic term * women | -9.83 (-16.92, -2.74) | 0.007 | -1.98 (-7.09, 3.14) | 0.448 | 0.36 (-2.13, 2.86) | 0.774 |
| Spline 2, cubic term * women | 0.90 (0.24, 1.55) | 0.007 | 0.55 (-1.83, 2.93) | 0.650 | -0.04 (-0.32, 0.23) | 0.758 |
| Intercept | -5.25 (-5.77, -4.72) | <0.001 | -5.87 (-6.20, -5.54) | <0.001 | -4.00 (-4.24, -3.76) | <0.001 |
| Intercept variance | 4.79 (4.20, 5.47) | | 7.06 (6.39, 7.80) | | 6.67 (6.13, 7.26) | |

*Note.* Unadjusted results. BCS70: 1970 British Cohort Study; NCDS: 1958 National Child Development Study; NSHD: 1946 National Survey of Health and Development.

**eTable 10.2. Marginal mean levels predicted from the multilevel growth curve models with caseness as outcome (logistic models)**

| | Age | Year | Marginal mean (95% CI) |
|---|---|---|---|
| NSHD | 36 | 1982 | 0.06 (0.05, 0.07) |
| | 43 | 1989 | 0.11 (0.10, 0.12) |
| | 53 | 1999 | 0.16 (0.15, 0.18) |
| | 63 | 2009 | 0.14 (0.13, 0.16) |
| | 69 | 2015 | 0.10 (0.09, 0.12) |
| | 74 | May 2020 | 0.30 (0.27, 0.32) |
| | 74.5 | Sept/Oct 2020 | 0.30 (0.26, 0.35) |
| | 75 | Feb/Mar 2021 | 0.27 (0.22, 0.31) |
| | **Age** | **Year** | **Marginal mean (95% CI)** |
| NCDS | 23 | 1981 | 0.08 (0.07, 0.08) |
| | 33 | 1991 | 0.06 (0.06, 0.07) |
| | 42 | 2000 | 0.12 (0.11, 0.12) |
| | 50 | 2008 | 0.14 (0.13, 0.15) |
| | 61.7 | Jan/Mar 2020 | 0.13 (0.12, 0.15) |
| | 62 | May 2020 | 0.12 (0.11, 0.13) |
| | 62.5 | Sept/Oct 2020 | 0.16 (0.15, 0.17) |
| | 63 | Feb/Mar 2021 | 0.15 (0.14, 0.16) |
| | **Age** | **Year** | **Marginal mean (95% CI)** |
| BCS70 | 26 | 1996 | 0.15 (0.14, 0.16) |
| | 29 | 1999 | 0.13 (0.13, 0.14) |
| | 34 | 2004 | 0.14 (0.13, 0.14) |
| | 42 | 2012 | 0.18 (0.17, 0.19) |
| | 46 | 2016 | 0.19 (0.18, 0.20) |
| | 50 | May 2020 | 0.18 (0.17, 0.19) |
| | 50.5 | Sept/Oct 2020 | 0.22 (0.21, 0.23) |
| | 51 | Feb/Mar 2021 | 0.20 (0.19, 0.22) |

*Note.* Unadjusted results. BCS70: 1970 British Cohort Study; NCDS: 1958 National Child Development Study; NSHD: 1946 National Survey of Health and Development.

**eFigure 10.1. Marginal predicted mean probability of psychological distress over time (year and age)**

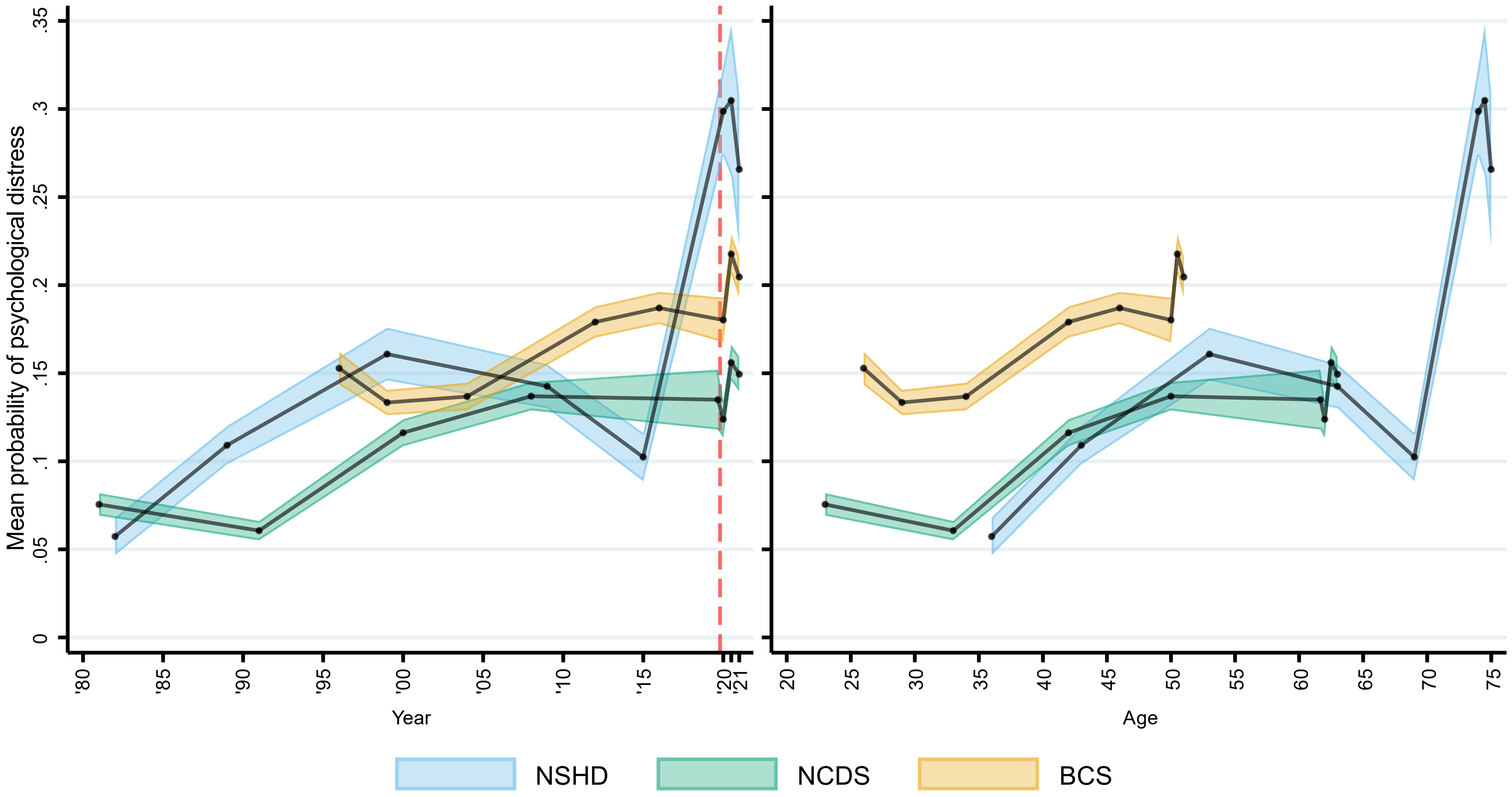

Note: Unadjusted results . 95% confidence intervals are indicated in lighter shaded areasBCS70: 1970 British Cohort Study, NCDS: 1958 National Child and Development Study; NSHD: 1946 National Survey of Health and Development .
The dashed line represents the first nationwide lockdown enforced in March 2020.

### eAppendix 11. Results from multilevel growth curve models with factor scores as outcome (linear models), 7 harmonised psychological distress indicators in NSHD

**eTable 11.1. Model coefficients from the multilevel growth curve models with factor scores as outcome (linear models), 7 harmonised indicators in NSHD**

| Models without interaction by birth sex | Coefficient (95% CI) | *p* |
|---|---|---|
| Spline 1, linear term | 0.05 (0.04, 0.07) | <0.001 |
| Spline 1, quadratic term | -0.002 (-0.002, -0.001) | <0.001 |
| Spline 2, linear term | -2.17 (-9.18, 4.83) | 0.543 |
| Spline 2, quadratic term | 0.84 (-1.76, 3.43) | 0.527 |
| Spline 2, cubic term | -0.08 (-0.31, 0.16) | 0.530 |
| Intercept | -0.25 (-0.34, -0.16) | <0.001 |
| Intercept variance | 1.23 (1.12, 1.35) | |

| Models with interaction by birth sex | Coefficient (95% CI) | *p* |
|---|---|---|
| Spline 1, linear term | 0.05 (0.04, 0.07) | <0.001 |
| Spline 1, quadratic term | -0.002 (-0.002, -0.001) | <0.001 |
| Spline 2, linear term | -8.22 (-14.46, -1.97) | 0.010 |
| Spline 2, quadratic term | 3.04 (0.74, 5.35) | 0.010 |
| Spline 2, cubic term | -0.28 (-0.49, -0.07) | 0.010 |
| Intercept * women | 0.40 (0.22, 0.57) | <0.001 |
| Spline 1, linear term * women | 0.00 (-0.03, 0.03) | 0.943 |
| Spline 1, quadratic term * women | 0.000 (-0.001, 0.001) | 0.860 |
| Spline 2, linear term * women | 12.06 (-1.41, 25.53) | 0.079 |
| Spline 2, quadratic term * women | -4.41 (-9.39, 0.58) | 0.083 |
| Spline 2, cubic term * women | 0.40 (-0.06, 0.86) | 0.086 |
| Intercept | -0.46 (-0.58, -0.34) | <0.001 |
| Intercept variance | 1.17 (1.06, 1.29) | |

*Note.* Unadjusted results.

**eTable 11.2. Marginal mean levels predicted from the multilevel growth curve models with factor scores as outcome (linear models), 7 harmonised indicators in NSHD**

| Age | Year | Marginal mean (95% CI) |
|---|---|---|
| 36 | 1982 | -0.25 (-0.34, -0.16) |
| 43 | 1989 | 0.05 (-0.02, 0.12) |
| 53 | 1999 | 0.17 (0.08, 0.25) |
| 63 | 2009 | -0.07 (-0.14, 0.01) |
| 69 | 2015 | -0.38 (-0.47, -0.29) |
| 74 | May 2020 | 0.09 (-0.03, 0.22) |
| 74.5 | Sept/Oct 2020 | 0.23 (-0.01, 0.47) |
| 75 | Feb/Mar 2021 | 0.16 (-0.09, 0.40) |

*Note.* Unadjusted results.

**eFigure 11.1. Marginal mean psychological distress factor scores over time (year), 7 harmonised psychological distress indicators in NSHD**

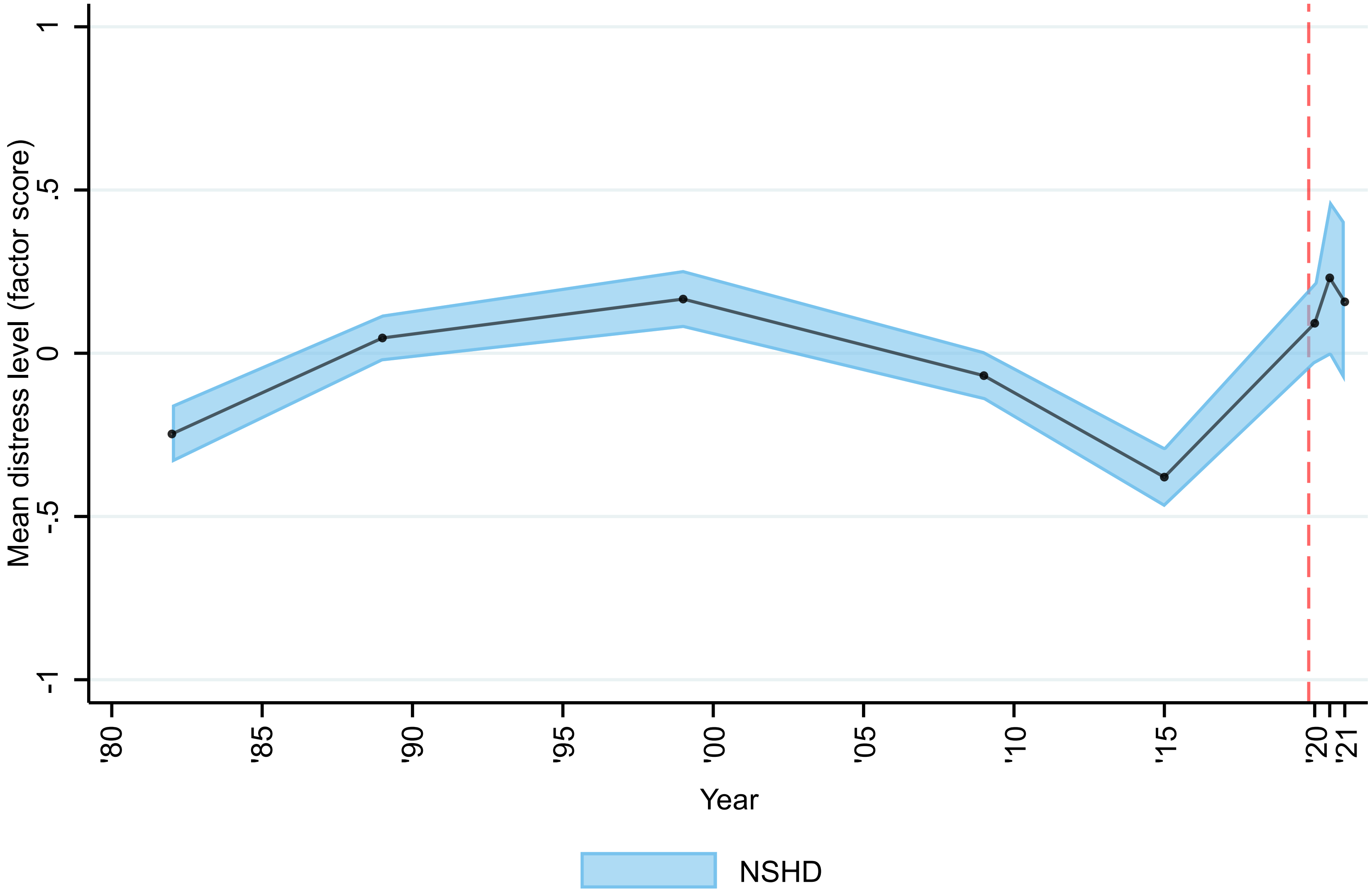

Note: Unadjusted results. 95% confidence intervals are indicated in lighter shaded areas. NSHD: 1946 National Survey of Health and Development. The dashed line represents the first nationwide lockdown enforced in March 2020.
